# Social inequalities in vaccine coverage and their effects on epidemic spreading

**DOI:** 10.1101/2024.11.01.24316556

**Authors:** Adriana Manna, Márton Karsai, Nicola Perra

## Abstract

Vaccinations are fundamental public health interventions. Yet, inequalities in vaccines uptake across socioeconomic groups can significantly undermine their impact. Moreover, heterogeneities in vaccination coverage across socioeconomic strata are typically neglected by epidemic models and considered, if at all, only at posteriori. This limitation reduces their ability to predict and assess the effectiveness of vaccination campaigns. Here, we study the impact of socioeconomic inequalities in vaccination uptake on epidemic burden. We consider a modeling framework based on generalized contact matrices that extend traditional age-stratified approaches to incorporate socioeconomic status (SES) variables. We simulate epidemic dynamics under two scenarios. In the first vaccination campaigns are concurrent with epidemics. In the second instead, vaccinations are completed before the onset of infection waves. By using both synthetic and empirical generalized contact matrices, we find that inequalities in vaccine uptake can lead to non-linear effects on disease outcomes and exacerbate disease burden in disadvantaged groups of the population. We demonstrate that simple models ignoring SES heterogeneity produce incomplete or biased predictions of epidemic burden. Additionally, we show how inequalities in vaccine coverage interact with non-pharmaceutical interventions (NPIs) compounding differences across subgroups. Overall, our findings highlight the importance of integrating SES dimensions, alongside age, into epidemic models to inform more equitable and effective public health interventions and vaccination strategies.

## Introduction

Vaccination is one of the most effective public health interventions for controlling infectious diseases, protecting individuals and communities [1]. Besides their efficacy to different endpoints (e.g., infection, death), vaccine access and uptake are key factors shaping the impact on disease burden. Vaccines are designed to be accessible to all, but limited stockpiles often raise the issue of optimal distribution [2, 3]. Common protocols priorities essential workers, the elderly, children, or individuals with underlying health conditions. Social status should not determine access and yet, as clearly observed during the COVID-19 Pandemic, vaccination access is shaped by significant inequalities across and within countries [4–9]. Indeed, studies have shown that individuals from lower socioeconomic backgrounds are less likely to be vaccinated against COVID-19 [10–12]. For example, disparities have been observed in Florida’s initial vaccine rollout [13], where lower-income and minority communities faced significant obstacles in accessing vaccines. Geographic and ethnic disparities also play a crucial role, as seen in Hungary, where socioeconomic deprivation has been found to be correlated with lower vaccination coverage [12, 14, 15]. In the Greater Manchester area in the UK, COVID-19 vaccination rates have been significantly lower among Black and Asian ethnic groups compared to others [16].

The correlation between socioeconomic status (SES) and vaccination uptake varies across different illnesses [17], but individuals experiencing lower SES exhibit lower vaccination rates due to barriers like limited access to healthcare, but also exposure to (mis)information, and vaccine hesitancy [18]. Indeed, social and behavioural factors emergent from interactions and (mis)information exposure play an important role in vaccine uptake [19, 20]. Studies in France have highlighted higher COVID-19 vaccine hesitancy among groups experiencing lower SES, driven by concerns over vaccine safety and distrust in government health measures [21, 22]. Overall, these findings remain consistent regardless of age, indicating that socioeconomic factors play a crucial role in vaccine hesitancy and access across all age groups [23, 24].

Additionally, for any given uptake and coverage level, the impact of a vaccination campaign is closely tied to its timing and speed relative to the progression of the epidemic. A campaign that begins only after a large portion of the population has already been infected will inevitably have a much smaller effect compared to one initiated earlier. During the COVID-19 Pandemic, vaccinations started in December 2020 (in the Global North) [25, 26]. At that time many countries were facing a rise in infections induced by the winter seasonality and by the spread of a more transmissible variant (i.e., Alpha). Furthermore, doses were a scarce resource and countries had to deal with tremendous logistical challenges for their distribution. As a result, the initial vaccination rates were far from optimal. Hence, the beginning of COVID-19 vaccinations is a clear example of a campaign that starts amid an epidemic that instead finds a large majority of the population susceptible. Non-pharmaceutical interventions (NPIs) have been critical to support these complex phases. However, NPIs have also been associated with socioeconomic inequalities. Indeed, the ability to self-isolate, work from home, and avoid crowded places varied significantly across different socioeconomic groups [9, 27–31]. As a result, disadvantaged groups of the population suffered from compounding inequalities. On one side, they had limited access to vaccines. On the other, they could not afford to reduce the risk of infection by adopting NPIs. These disparities have significant implications for public health, leading to sub-optimal vaccination coverage and perpetuating the spread of vaccine-preventable diseases.

In this context, incorporating socioeconomic factors in epidemic models beyond the traditional age-wise stratification is essential [32–37]. However, current approaches fail to do so potentially resulting in incomplete or biased predictions about epidemic burden and vaccination campaign effectiveness. To address these complexities and limitations, here we adopt and extend the epidemic framework proposed in Ref. [38] that features *generalized contact matrices*. The model allows accounting for the stratification of contacts across multiple dimensions such as age and SES [38]. In doing so, it provides a finer description of the population under investigation and allows to assess how disparities in vaccination coverage and NPIs adoption influence the overall epidemic outcome. To isolate the interplay between social contacts and vaccination uptake across subgroups we consider both synthetic and real generalized contact matrices that stratify contacts according to age and one SES indicator. The age stratification is data-driven in both cases, while the stratification for the SES dimension is built either by using a simple model or from data collected in Hungary [39, 40]. To account for the effect of vaccination timing, these analyses are developed in two distinct scenarios. The first simulates a situation where the vaccination campaign and the epidemic are concurrent. The second scenario instead simulates a situation in which the vaccination campaign begins before the epidemic, reaching a substantial portion of the population prior to the outbreak.

Our results emphasize the importance of models that explicitly account for socioeconomic disparities to accurately estimate the epidemic burden within specific subgroups of the population. Indeed, we demonstrate that, when disparities in vaccination uptake exist across subgroups, models that do not incorporate these dimensions fail to accurately predict the epidemic burden within these groups. Furthermore, we find that the unequal distribution of vaccines across socioeconomic groups might lead to a non-linear effect on epidemic outcomes that, in general, cannot be estimated a posteriori from models that account just for the traditional age stratification. Finally, our findings show how disparities intensify when compounded by the interaction with NPIs.

Overall, our results underscore the importance of incorporating subgroup-specific factors into epidemic models to enhance the accuracy of predictions and, in turn, the effectiveness of public health interventions and vaccination strategies.

## Results

We consider a Susceptible-Exposed-Infectious-Recovered (SEIR) compartmental model incorporating multidimensional contact matrices with an additional dimension with three levels (i.e., subgroups), besides age [38]. We take into account eight age classes [0 − 5), [5 − 15), [15 − 30), [30 − 45), [45 − 60), [60 − 70), [70 − 80), [80+), and three SES: low, middle, and high. Specifically, we use *generalized contact matrices* **G** as multidimensional objects, whose elements *G*_**a**,**b**_ capture the contact rates between individuals in group **a** and **b**. Here, **a** = (*i, α*) and **b** = (*j, β*) are index vectors (i.e., tuples) representing individual’s membership to each category defined along age *i, j* and the second dimension *α, β*. The stratification for age is obtained from data [39, 40], while the stratification for SES is either synthetic, obtained from a simple model, or measured from data. In the first case, we introduce assortativity by assuming that 60%, 50%, and 65% of the contacts in the first, second, and third SES category take place within each group. Furthermore, we assume that activity levels are distributed heterogeneously, with 20%, 40%, and 40% of the total contacts allocated to the first, second, and third groups, respectively. We set an uneven population distribution with group sizes comprising 35%, 45%, and 20% of the total population. The model used to generate the synthetic population and mixing patterns along the second dimension follows Ref. [38].

We model a vaccination rollout, where each day a fraction of the population receives a vaccine that reduces the probability of infection and death by 60% and 80% respectively [41]. Additionally, we assume that infected vaccinated individuals are 40% less likely to transmit the virus further. The daily allocation of vaccines to age groups is taken from data in Hungary reflecting the COVID-19 vaccination campaign (See SI Section 1.1). We study and compare three different vaccination uptake distributions (VD for short) across the SES subgroups. In the first case (*V D*1), each subgroup receives an equal share, i.e., one-third of the vaccines. In the second case (*V D*2), the second subgroup (middle SES) receives 40% of the available vaccines each day, while the first (low SES) and third (high SES) subgroups receive each 30%. Finally, in the third case (*V D*3), the distribution is more uneven, especially for the subgroups experiencing a low SES, with the three subgroups receiving 25%, 35%, and 40% of the vaccines, respectively. Under all vaccination distributions, we assume that each subgroup can be vaccinated up to 95%. We refer the reader to the Materials and Methods section and the Supplementary Information for more details about the model and simulation setup.

Finally, as mentioned in the introduction, the interaction between the timing of a vaccination campaign and the progression of the epidemic plays a critical role in shaping the epidemic burden. To investigate their effect on the epidemic outcome, we focus on two simulation scenarios. In the first (*Scenario 1*), the epidemic and vaccination campaign start simultaneously. We set the basic reproduction number *R*_0_, defined as the number of secondary infections caused by a single infected individual in an otherwise fully susceptible population [42], to 1.6, representing a relatively slow epidemic and leading to a substantial overlap between the epidemic wave and the vaccination campaign. In the second scenario,(*Scenario 2*), the vaccination campaign is completed before the epidemic begins. Here, contrary to the previous scenario, we set a relatively high value of the basic reproductive number, *R*_0_ = 3, to simulate more rapid epidemic dynamics. The higher *R*_0_ allows us to observe the rapid spread of the virus in a predominantly vaccinated population.

### Scenario 1: concurrent vaccinations and epidemics

In Figure 1*a* we begin by observing the epidemic wave under different vaccination distributions (i.e., *V D*1, *V D*2, *V D*3) as predicted by the generalized model (see coloured solid lines) and an age-stratified model (see black dashed line). The first observation is that, as expected from Ref. [38], the generalised model’s projections significantly differ from those of the age-stratified model. Indeed, the latter overestimates the total number of infected individuals. The discrepancy is due to the distribution of activity, assortativity and population among subgroups. Second, the total number of infected projected by the generalized model varies across the vaccination uptake distributions. Notably, vaccination distribution of *V D*3 (in orange) proves to be the most effective, as it reduces the number of infections the most. This effectiveness is primarily due to the allocation of doses, where 75% of the total daily available vaccines are distributed to the two most socially active groups. Specifically, the third group, which, despite being the smallest by population size, exhibits the highest assortativity and activity (alongside the second group), receives 40% of doses. The combination of being both the smallest and the most active means that, in *V D*3, the third group becomes entirely vaccinated (up to the saturation value).

**Figure 1:**
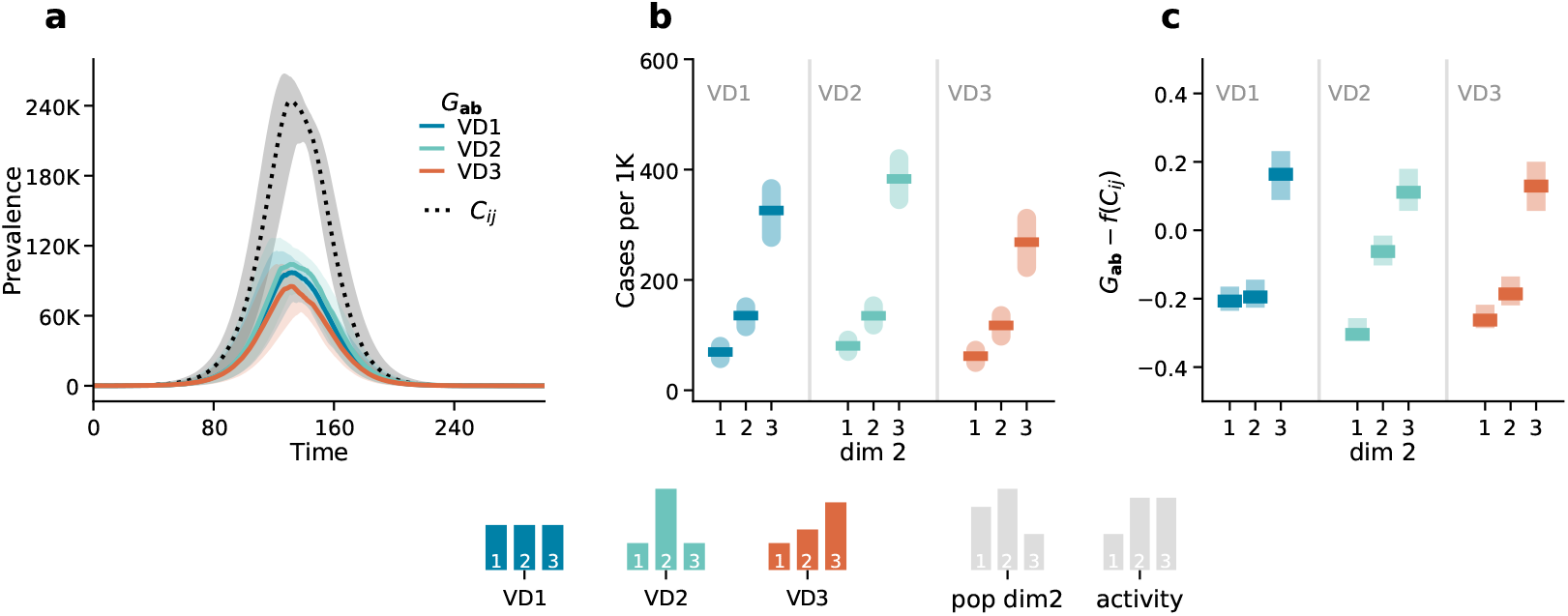
Scenario 1: epidemic outcomes. Panel *a* displays the number of infected individuals over time. The dotted black line corresponds to the outcome of the age-stratified model (*C*_*ij*_), and the solid coloured lines correspond to the outcomes of the generalized model (*G*_**ab**_) in the three different vaccination strategies (*V D*1, *V D*2, *V D*3). Panel *b* shows the attack rate by the second dimension (dim 2) predicted by the generalized model (*G*_**ab**_. Panel *c* shows the difference between the attack rates predicted by the generalized model *G*_**ab**_ and those estimated from the aggregate output of the age-stratified model *f* (*C*_*ij*_ by the second dimension (dim 2). Results refer to the median of 500 runs with IQRs. Epidemiological parameters: Γ = 0.25, Ψ = 0.4, *g*_1_ = 0.6, *R*_0_ = 1.6. Simulations start with *I*_0_ = 100 initial infectious seeds.

At the same time the second group, also highly active, receives 35% of doses. Prioritizing these two active groups maximizes the impact of the vaccination campaign. On the contrary, a different outcome is produced by *V D*2 (in green). In this case, 40% of the vaccines are allocated to the second group, which is both the most active (along with group 3) and the largest in population size. However, the vaccination share allocated to the second group is insufficient to effectively hamper infections. Simultaneously, the lower vaccination rate in the third group, combined with its high activity level, allows for increased transmission rates, which affect the overall dynamics of the epidemic. Similar results are observed for *V D*1 (in blue). Although doses are distributed evenly, the unequal population size and activity levels result in a performance comparable to *V D*2.

The generalized model allows us to estimate the disease burden across the three socioeconomic subgroups by summing over all age groups. For example, to compute the total number of infected individuals in subgroup *α* at time *t*, we can sum over all age groups i.e., *I*_*α*_(*t*) = ∑_*i*_ *I*_*i,α*_(*t*). Exploiting this feature of the generalized model, in panel *b* of Fig. 1, we examine the attack rates predicted by the generalised model by subgroup for each vaccination distribution. Interestingly, the third group consistently appears as the most affected, while the first group remains the least infected across all vaccination distributions. This is solely due to the differences in activity levels: as the third group is the most active, it is always the most infected relative to its population size. Notably, in *V D*2, the disparity in infections for the third group is even more pronounced, as it receives significantly fewer vaccines compared to the other groups, further amplifying its vulnerability.

Finally, we investigate whether, and to what extent, it is possible to predict these outcomes from models that do not explicitly account for SES. First, we point out that infections across subgroups of the population, not explicitly considered by a model, can be estimated only a posteriori by leveraging outputs of the age-stratified model, such as attack rates for vaccinated (*AR*_*V*_) and non-vaccinated (*AR*_*NV*_) individuals. We developed a technique to estimate the number of infections within each subgroup *α* by multiplying the attack rates by the total number of vaccinated and non-vaccinated individuals in each subgroup at time *t*. For further details on this method, we refer the reader to the Methods section and Supplementary Information. This technique has two significant limitations: 1) it assumes precise knowledge of the number of vaccinated individuals in each subgroup defined by age and SES, which might not be realistic in practice, 2) it cannot, by definition, capture potentially different dynamics such as different peak times across subgroups unseen by the model.

In panel *c* of Fig.1 we show the difference between the number of cases in each subgroup projected by the generalized model *G*_**ab**_ and estimated from an age-stratified model *f* (*C*_*ij*_)^1^. The differences between the two are consistently away from zero. Indeed, the epidemic burden within each subgroup results from the interplay among activity levels, contact patterns, population distribution, and vaccination coverage. This cannot be captured by traditional models that do not account, explicitly, for these factors. In Section 6 of Supplementary Information we further explore these results by running the same analysis under different vaccination distribution settings and a homogeneous mixing case. Overall, we observe the same qualitative findings.

#### The interplay between vaccinations and NPIs

We now investigate how the interplay between heterogeneities in adherence to NPIs and vaccination distribution among subgroups affects epidemic outcomes. We explore hypothetical cases where a population modifies its behaviour in response to the introduction of NPIs during an ongoing vaccination campaign. We assume that, to reduce contacts, NPIs are introduced 65 days after the onset of the epidemic leading to a 20% overall reduction in contact rates, though with variations across subgroups. Indeed, we assume that the ability to adhere to the NPIs is associated with membership in a particular population group. Specifically, we explore a case where people in the first group (i.e., experiencing the lowest SES) cannot afford to protect themselves as the other two groups. We assume that the NPIs introduce changes in both assortativity and activity. Specifically, due to the NPIs, 50%, 60%, and 70% of the contacts for the first, second, and third groups, respectively, take place within their group. Thus, assortativity increases across all groups, with the second and third groups experiencing the most significant rise. Additionally, NPIs shift activity levels to 40%, 25%, and 35% for the first, second, and third groups, respectively. This results in a decrease in activity for the second and third groups, while the first group experiences a relative increase, further exacerbating its vulnerability. Following the same logic as before, we study the effects of three different vaccination uptake distributions (i.e., *V D*1, *V D*2, *V D*3).

In Fig. 2*a*, we report the results in a baseline case where the disease spreads unmitigated by any NPIs. The first column of panel Fig. 2*a* displays the prevalence over time, while the second column shows the corresponding attack rates per 1000. The differences between all vaccination strategies are small, but *V D*3 is still the most effective, leading to a higher reduction. In contrast, *V D*1 and *V D*2 are less effective, with *V D*2 being the least effective.

**Figure 2:**
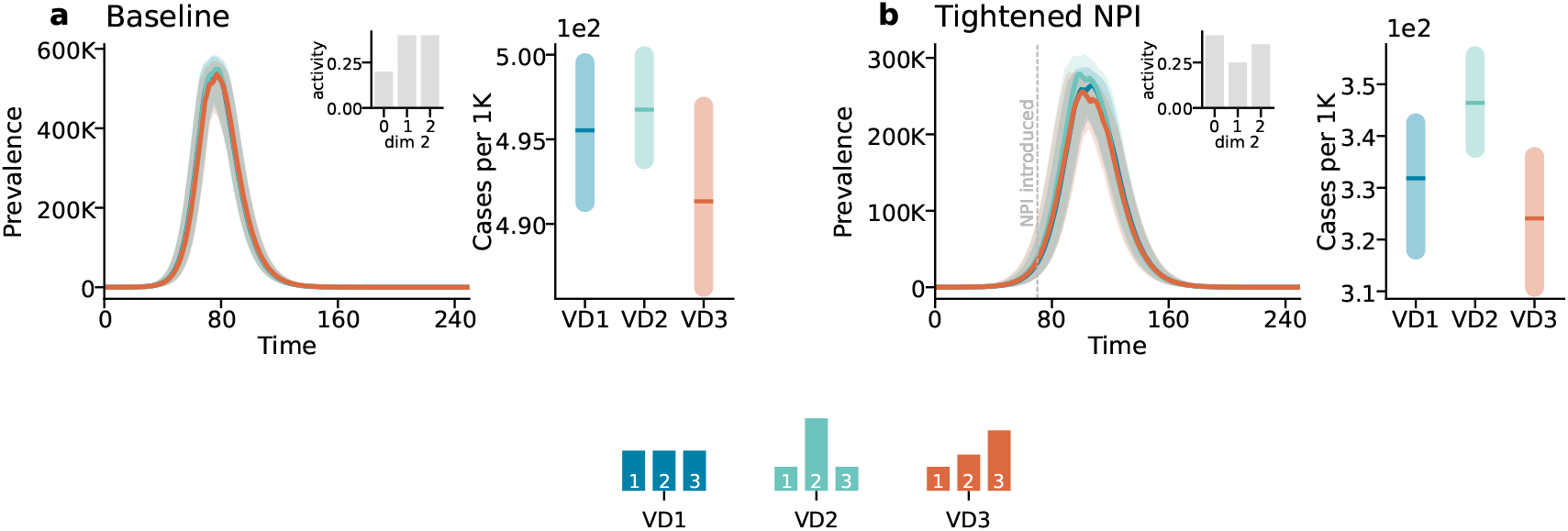
Scenario 1: The impact of NPIs under different vaccination distributions. Panel *a* refers to the baseline case, with insets depicting the activity distribution along the second dimension of the generalised contact matrix. The first column shows the prevalence, while the second column the attack rates per 1000. Panel *b* follows the same structure but refers to the case where NPIs are introduced, reducing contacts by 20%. The epidemic starts at *t*_*epi*_ = 0 with the onset of vaccination. NPIs are introduced 65 days after the epidemic onset. Results represent the median of 500 runs with confidence intervals. Epidemiological parameters: Γ = 0.25, Ψ = 0.4, *g*_1_ = 0.5, *g*_2_ = 0.8, and *R*_0_ = 2.5. Simulations begin with *I*_0_ = 100 initial infected seeds.

However, as shown in Fig. 2*b*, when NPIs are introduced, the attack rate of the different SES changes (See SI Section 7 for the corresponding plot) thus influencing the relative effectiveness of the vaccination strategies. By ensuring better coverage for the first group, which is now more vulnerable due to its higher activity, strategy *V D*1 results in a more balanced reduction of cases across all subgroups. Meanwhile, *V D*2 becomes even less effective, as the majority of vaccines are assigned to the second group, the less active, while the more active groups receive fewer vaccines. Additionally, *V D*3 loses its relative efficacy due to the reduced activity of group three, which receives the highest share of vaccines, and the increased activity in group one, which is the least vaccinated. For completeness, in the Supplementary Information, we investigate the effect of mortality by modelling the daily number of deaths. In SI Section 7 we report the corresponding results, which show the same trend.

### Scenario 2: vaccinations completed before epidemics

In this scenario, we explore the epidemic dynamics assuming the vaccination rollout is already completed before the onset of the epidemic. This represents situations akin to a post-vaccination wave of infection, where the majority of the population has received vaccines, but a virus continues to circulate, possibly due to new variants or waning immunity.

Following the structure of the previous scenario, Fig. 3*a* illustrates the number of newly infected individuals over time for the three vaccination distributions, as projected by the generalized model (see coloured solid lines) and the traditional age-stratified model (see black dotted line). Also in this scenario, the generalized model projects different epidemic outcomes compared to the age-stratified model. Additionally, the differences between epidemic outcomes across vaccination distributions are more pronounced. This is because the epidemic starts only after the vaccinations have been fully implemented, maximizing the impact of the different vaccination strategies. Notably, *V D*3 remains the most effective strategy, resulting in a significantly lower number of infections. The success of *V D*3 stems from the priority given to the most active groups, ensuring that the most socially active members of the population are well-protected. Conversely, *V D*2, which prioritizes the middle SES group, leads to a higher overall number of cases, as the third, more active SES group remains under-vaccinated and plays a critical role in sustaining transmission.

**Figure 3:**
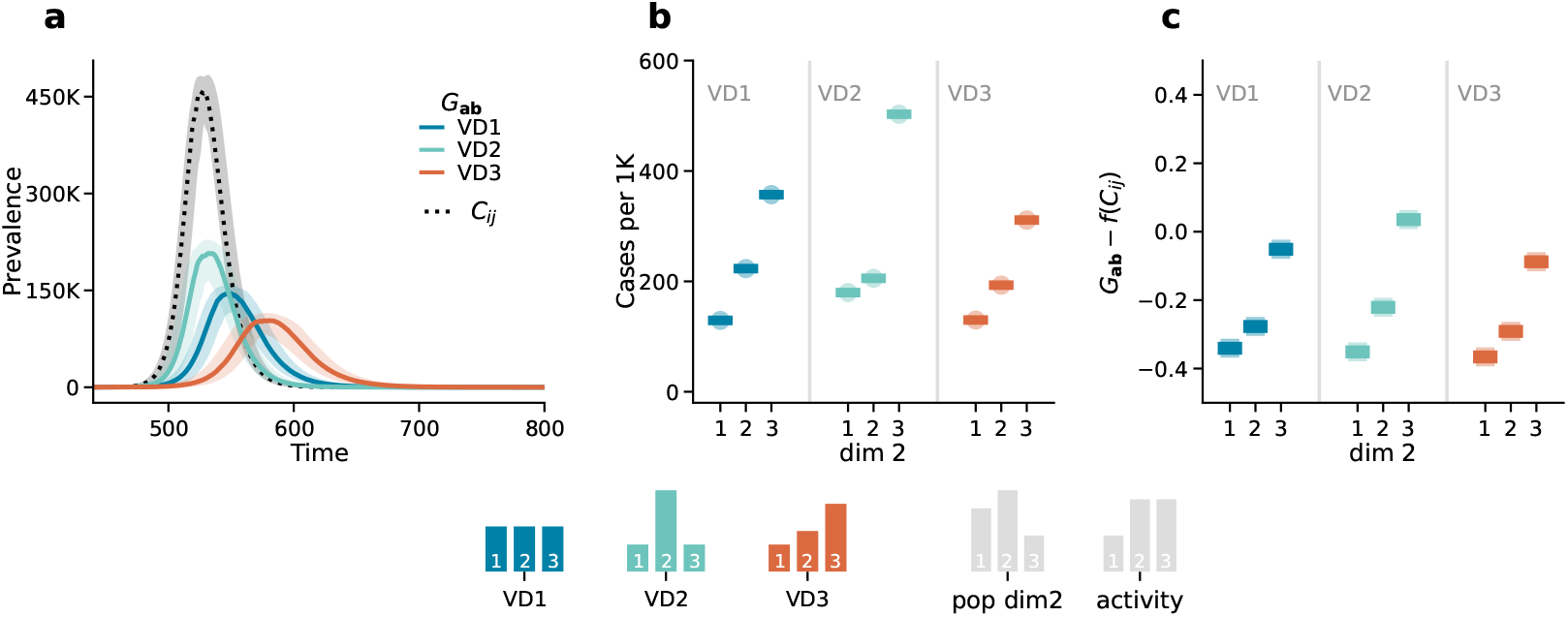
Scenario 2. Epidemic outcomes: Panel *a* displays the prevalence over time. The dotted black line corresponds to the outcome of the age-stratified model (*C*_*ij*_), and the solid coloured lines correspond to the outcomes of the generalized model (*G*_**ab**_) in the three different vaccination distribution strategies (i.e., *V D*1, *V D*2, *V D*3). Panel *b* shows the attack rates per 1000 by the second dimension (dim 2) projected by the generalized model (*G*_**ab**_. Panel *c* shows the difference between the attack rates projected by the generalized model *G*_**ab**_ and those estimated from the aggregate output of the age-stratified model *f* (*C*_*ij*_ by the second dimension (dim 2). Results refer to the median of 500 runs with IQRs. Epidemiological parameters: Γ = 0.25, Ψ = 0.4, *g*_1_ = 0.6, *R*_0_ = 3. Simulations start with *I*_0_ = 100 initial infectious seeds.

Fig. 3*b* breaks down the attack rates by SES, showing that even in this scenario, the most active group (SES 3) experiences the highest number of infections relative to their population size, particularly in *V D*2. This confirms that, despite overall high vaccination coverage, specific subgroups can still disproportionately contribute to the spread of the virus.

In panel 2*c*, we compare the predictions of the generalized model (*G*_**ab**_) with those derived from the age-stratified model (*C*_*ij*_) using the same technique discussed above (See MM and SI Section 4). Once again, we observe significant discrepancies between the methods. Overall, these results highlight the importance of explicitly incorporating subgroup-specific dimensions, such as socioeconomic status, into epidemic models.

In the Supplementary Information, we further explore these results by conducting a robustness analysis running the same analysis under a different vaccination distribution settings. In particular, we consider a case which age is ignored and another that assumes homogeneous mixing between groups. Notably, in the latter, we find that the difference between the projected burden of the epidemic from aggregate model output (*f* (*C*_*ij*_)) and those gathered from the generalized model(*G*_**ab**_) is consistently 0. This implies that in case of homogeneous mixing and when the overlap between vaccination and epidemics is zero the differences across sub-groups can be obtained also a posteriori from models that neglected them.

#### The interplay between vaccinations and NPIs

In this scenario, the population is already fully vaccinated before the start of the epidemics. We assume that, before the start of the vaccination, some NPIs were in place. Hence we consider a situation where NPIs are then relaxed, leading to a 20% overall increase in contact rates. As before, we assume that the ability to adhere to NPIs varies across subgroups, with the variations remaining consistent with those depicted in Fig. 2. The NPIs are reduced 65 days after the onset of the epidemic.

In the baseline case (see Fig. 4*a*), although the different impact of vaccination distributions is more pronounced compared to the previous case, the relative effectiveness of the vaccination strategies follows the order *V D*2, *V D*1, and *V D*3, with *V D*2 being the least effective. However, when NPIs are relaxed (see Fig. 4*b*), the epidemic burden in the different SES changes according to the new activity distribution, and in turn affects the relative effectiveness of the strategies. In this case, strategy *V D*1 becomes more effective than *V D*3, while *V D*2 remains the least effective vaccination strategy.

**Figure 4:**
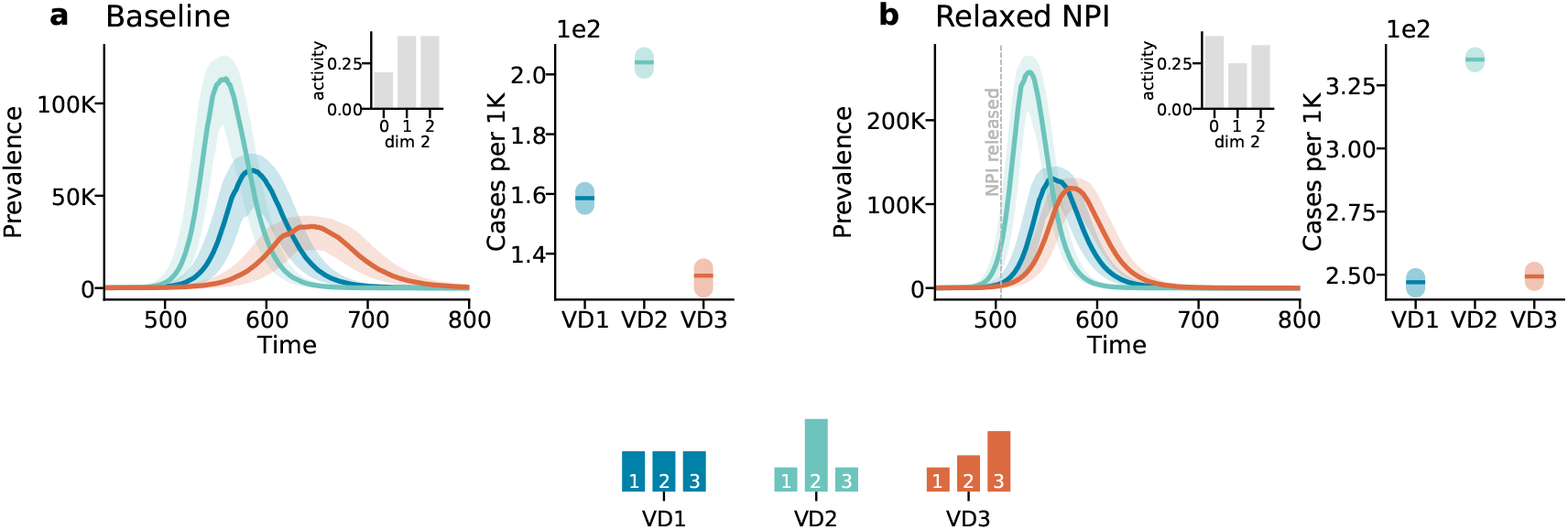
Scenario 2: The impact of NPIs under different vaccination distributions. Panel *a* refers to the baseline case, with insets depicting the activity distribution along the second dimension. The first column shows the prevalence as function of time, while the second column shows the attack rate scaled by 1000. Panel *b* follows the same structure but refers to the case where NPIs are released, increasing contacts by 20%. NPIs are adjusted 65 days after the epidemic onset. Results represent the median of 500 runs with confidence intervals. Epidemiological parameters: Γ = 0.25, Ψ = 0.4, *g*_1_ = 0.5, *g*_2_ = 0.8, and *R*_0_ = 2.5. Simulations begin with *I*_0_ = 100 initial infected seeds.

These simulations highlight how the interplay between NPIs and vaccination strategies influences epidemic outcomes, with effectiveness depending on the timing and intensity of interventions. Again, in the Supplementary Information, we investigate the effect of mortality which shows the same trends (See SI Section 7 for the corresponding results).

### Hungarian contact data

We applied the model to empirical data describing social contacts stratified by age and one SES variable (i.e., self-perceived wealth with respect to the average) in Hungary during the COVID-19 Pandemic. The data has been collected via computer-assisted surveys from 1000 respondents describing a representative sample of the Hungarian adult population in terms of gender, age, education level, and type of settlement [39, 40]. To build these matrices we used two contact diaries collected in April and November 2021, respectively just before the third and the fourth wave of the COVID-19 Pandemic in Hungary. We refer the reader to the Materials and Methods section and the Supplementary Information for more details about the data and its collection.

In Fig. 5, we present the outcomes for the Hungarian case, respectively for Scenario 1 (panel *a*) and Scenario 2 (panel *b*). In each panel, we show the number of cases per 1000 individuals stratified by the non-vaccinated (*Non* − *vax*) and vaccinated (*V ax*) sub-populations and overall (*All*). To facilitate straightforward visual comparison between outcomes, the *y-axis* of the three subplots in each panel is set to span the same range. In other words, the difference between the maximum and minimum values on the *y-axis* is fixed across all subplots within the same panel.

**Figure 5:**
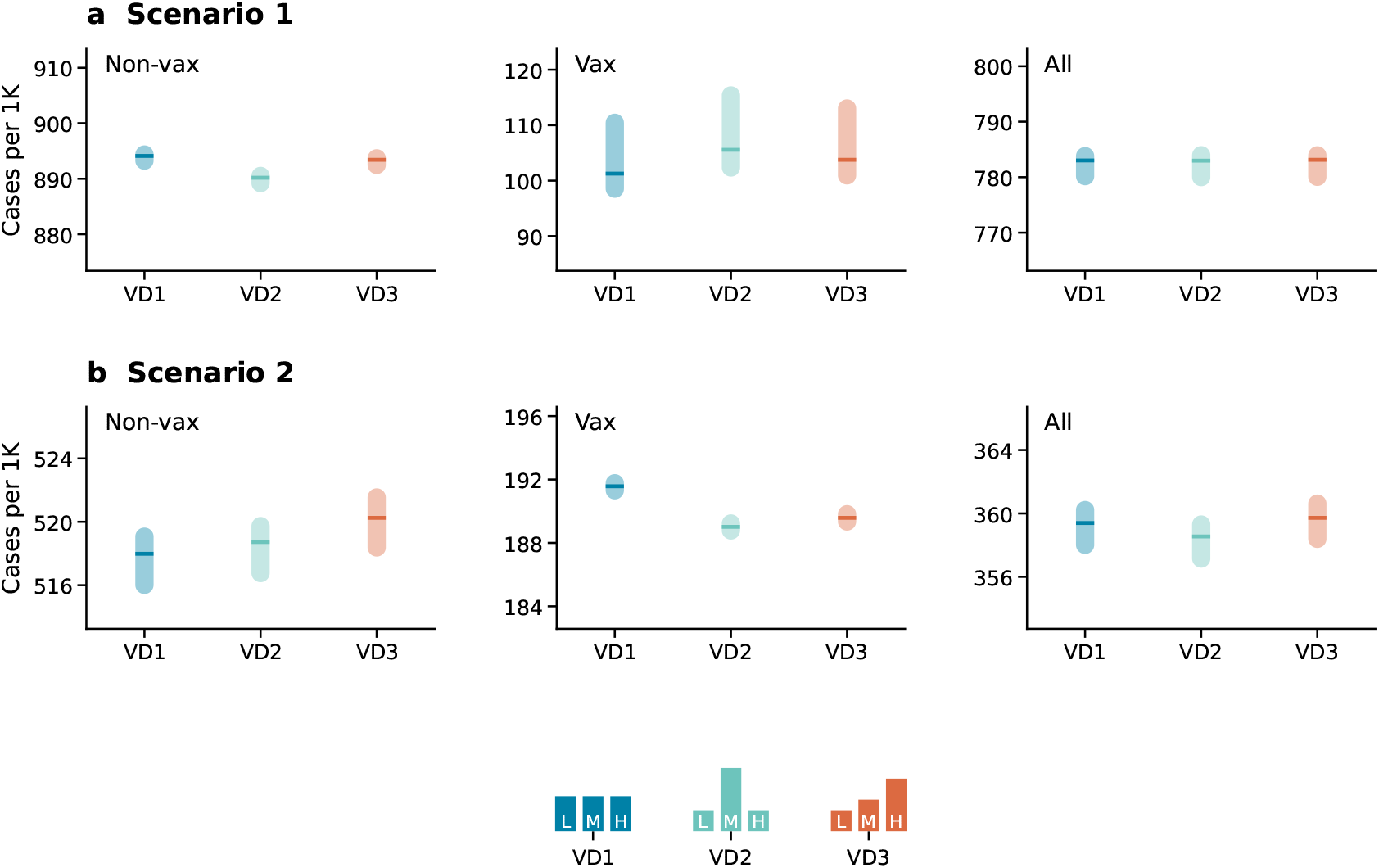
Epidemic outcomes under different vaccination distributions and real contact data. Panels in (*a*) refer to Scenario 1 and display the number of cases per 1000 stratified by vaccination status (non-vaccinated, vaccinated, and overall) and under different vaccination distributions (*V D*1, *V D*2, *V D*3). Panels *b* show the corresponding numbers for Scenario 2. Results refer to the median of 500 runs with confidence intervals. Epidemiological parameters: Γ = 0.25, Ψ = 0.4, *g*_1_ = 0.6, *g*_2_ = 0.8 and *R*_0_ = 3. Simulations start with *I*_0_ = 100 initial infectious seeds.

In Scenario 1, the population was initialized with 20% of individuals being immune, representing those who had residual immunity. Vaccination was assumed to have started simultaneously with the onset of the epidemic, setting the day of the epidemic as time *t*_epi_ = 0. For Scenario 2, the population was initialized with 40% of individuals being immune, reflecting a higher level of residual immunity. In this case, the epidemic was set to begin at *t*_epi_ = 220 when 55% of the population is vaccinated, which implies 220 days had passed since the start of the vaccination campaign before the epidemic started. This adjustment mirrors the same time gap observed between the start of the COVID-19 vaccination campaign and the onset of the fourth epidemic wave in Hungary.

The scenarios, modelled under different vaccination distributions (i.e., *V D*1, *V D*2, *V D*3), reveal distinct outcomes. In Scenario 1, the differences across vaccination distributions are small, as shown in Fig. 5*a*. This result can be explained by the fact that vaccination overlapped with a fairly fast epidemic. As a result, the effects of varying vaccination uptake are less visible, leading to similar outcomes.

In contrast, Scenario 2 (see Fig. 5*b*) shows more pronounced differences among the vaccination strategies. Here, *V D*2 appears to be the most effective in reducing the epidemic burden. Interestingly, as shown in the Supplementary Information, we find significant difference is also in the mortality of the non-vaccinated population, indicating that even non-vaccinated individuals benefit indirectly from effective vaccination strategies.

## Discussion

The COVID-19 Pandemic was a stark reminder that inequalities in vaccine accessibility and in non-pharmaceutical interventions compliance can shape the trajectory of an epidemic. Building on the mathematical framework of generalized contact matrices recently presented in Ref. [38], here we studied the impact of these inequalities.

The timing of vaccinations relative to epidemics is also a key factor. Hence, we explored two scenarios. In the first the epidemics and vaccinations begin simultaneously. In the second, the epidemics start in an almost fully vaccinated population. In these settings, we analyze the impact of different vaccination strategies, where subgroups of the population exhibit varying levels of vaccination uptake, and quantified the misrepresentation of simpler models, based only on age, that neglect heterogeneities along other dimensions.

Our results highlight the importance of using generalized models, that explicitly account for socioeconomic inequalities, to accurately estimate the epidemic burden within different subgroups of the population. Indeed, we compared the attack rates predicted by generalized models in each subgroup of the population (e.g., SES) with those inferred a posteriori from the outputs of age-stratified models. Interestingly, we found consistent differences between the two. Hence, disparities in vaccination uptake and contact patterns among subgroups make it challenging to estimate outcomes in subgroups of the population from models that do not consider these additional stratifications. To test these findings further, we conducted a sensitivity analysis by varying the values of *R*_0_ and the timing of the epidemics’ onset (*t*_epi_). Adjusting these two parameters modulates the overlap between epidemic waves and vaccinations, allowing us to explore situations in between the two scenarios. The analysis showed that, as we fix *R*_0_ and increase *t*_epi_ —thus reducing the overlap between epidemic waves and vaccinations— the difference between the two methods converges to a stable value, typically different than zero. A similar trend occurs when we increase *R*_0_ with *t*_epi_ set to zero. For further details, see Supplementary Information 6.2.2.

Additionally, we showed how NPIs, introduced or relaxed during an ongoing epidemic, can shift the relative effectiveness of vaccination strategies. Finally, using real generalized contact matrices from Hungary, we examined, in more realistic settings, how different vaccination distributions could influence the trajectory of an epidemic. Also the results obtained considering real contact patterns underscore the importance of incorporating socioeconomic factors into epidemic models.

It is important to acknowledge the limitations of our work. First, we note that our study relies solely on what-if scenarios designed to mimic real-world cases. However, we did not calibrate the model to epidemic data. The model used to generate synthetic generalized contact matrices, developed in Ref. [38], was not designed to replicate empirical data but to provide a flexible framework for exploration. The modelling of NPIs was guided by simplicity rather than realism. Indeed, our aim was to showcase the potential interactions between vaccination uptake, contact patterns, and NPIs, and to demonstrate their effects on the epidemic outcome, while providing a framework that can be adapted to different settings. Finally, a simplification was made in the modelling of the vaccination protocol by considering only a single dose that becomes immediately effective. More work is needed to address these limitations and extend the scope of the research presented here. To this end, modelling efforts should be assisted by progresses in data collection and sharing. Indeed, most of the relevant data is now disaggregated only by age thus limiting the scope and application of generalised models.

Overall, our study shows that incorporating socioeconomic heterogeneities into epidemic models allows for more accurate estimations of disease burden across different population subgroups. By accounting for inequalities in both contact patterns and vaccination uptake, our model provides a nuanced view of how these factors interact to influence epidemic dynamics. Our framework offers a straightforward, easy-to-implement, approach that enables fast and precise predictions of epidemic burden and vaccination coverage across different subgroups. This method could assist public health officials in tailoring vaccination campaigns and other interventions more effectively, ultimately reducing the unequal impact of epidemics on vulnerable populations.

## Materials and Methods

### Epidemiological model

We consider a Susceptible-Exposed-Infectious-Recovered (SEIR) compartmental model with vaccination where susceptible (*S*) are healthy individuals at risk of infection, exposed (*E*) are infected but not yet infectious, infectious (*I*) can spread the disease and recovered (*R*) are no longer infectious nor susceptible to the disease [43]. All compartments are then further stratified between non-vaccinated (NV) and vaccinated (V) individuals. In the Supporting Information we also model mortality by adding a death (*D*) compartment to the model.

We focus on two models that primarily differ in how they represent individual interactions: the age-stratified and the generalized model (See SI 2 for further details).

#### Age-stratified model

The age-stratified model uses contact matrices stratified by age **C** to account for the differences in contact patterns among various age groups. The element *C*_*ij*_ quantifies the average number of contacts that an individual in age-bracket *i* has with individuals in age group *j* within a certain time window [44–46]. The population is divided into age brackets so that 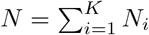. The variables *N*_*i*_ capture the number of individuals in age group *i* while *K* indicates the number of different age groups.

#### Generalized model

The generalized model extends the concept of age-stratified contact matrices to include multiple dimensions, represented by generalized contact matrices **G**. In more detail, we describe the generalized contact matrices as *G*_**a**,**b**_, where **a** = (*i, α*) and **b** = (*j, β*) are tuples (i.e., index vectors) representing individuals membership to each category. With these matrices we can, for example, capture contact stratification according to age and income (*α*). In this case, *G*_**a**,**b**_ would then describe the average number of contacts that an individual in age bracket *i* and income *α* has with people in age group *j* and income *β*, in a given time window. We refer the reader to the Supporting Information for more details.

#### Vaccinations

Vaccination data on the daily doses administered is available, at the lower level, by age groups (See SI Section 1.1) which we distribute among subgroups (SES) according to a given distribution *P*_*vax*_(*α*), which indicates the proportion of vaccines allocated to each subgroup. Let Ω_*i*_(*t*) represent the daily administered doses at time *t* for age group *i*. The number of vaccines administered to a subgroup *α* is then Ω_*i,α*_(*t*) = Ω_*i*_(*t*) · *P*_*vax*_(*α*). Each subgroup can be vaccinated up to 95%. If this limit is reached, any excess vaccinations are redistributed randomly within that particular age group.

#### Numerical simulations

We developed a stochastic, discrete-time, compartmental model where the transitions among compartments are simulated through chain binomial processes. In particular, at time step *t* the number of individuals in group **a** and compartment *X* transiting to compartment *Y* is sampled from *PrBin*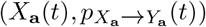, where 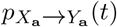 is the transition probability.

### Inferring epidemic burden for subgroups

To estimate the epidemic burden for specific subgroups (*α*) using outputs from a model stratified solely by age, we first need to infer the epidemic burden within each age group in the subgroup (*i, α*) and subsequently aggregate these age-specific results to obtain *AR*_*α*_. This requires estimating the number of susceptible individuals in each subgroup who became exposed during the epidemic i.e., 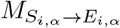. This involves calculating the attack rates for vaccinated *AR*_*V i*_ and non-vaccinated *AR*_*NV i*_ individuals, in each age group *i*, from the age-stratified model (See SI Section 3 for further details on this calculation). Therefore, we can multiply *AR*_*NV i*_ and *AR*_*V i*_ by the total number of non-vaccinated and vaccinated individuals in each group (*i, α*) at time *t*, taking into account the transitions between non-vaccinated and vaccinated states over time. The equation used for this calculation reads as follows:

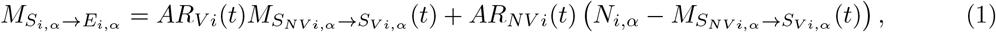

where *AR*_*V i*_(*t*) and *AR*_*NV i*_(*t*) represent respectively the attack rates for vaccinated and non-vaccinated individuals in age group *i* and 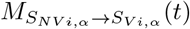 denotes the number of susceptible individuals in groups (*i, α*) who were vaccinated at time *t*. The attack rate for subgroups (*i, α*) can be then calculated by dividing equation (1) for the population size as follows:

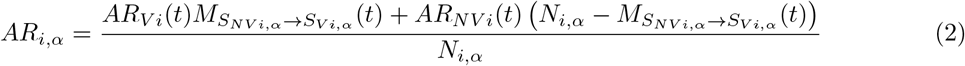

We can then aggregate by age groups equation (2) and divide by the total number of individuals in subgroup *α* as follows:

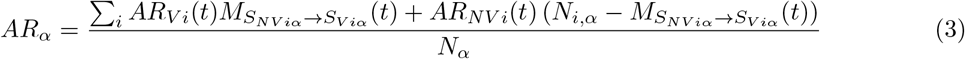

See the Supplementary Information 4 for further details.

### Real generalised contact matrices

The real generalised contact matrices used are obtained from the MASZK survey study [39, 40], a large data collection effort on social mixing patterns made during the COVID-19 pandemic, conducted in Hungary from April 2020 to July 2022. The study recorded 26 monthly cross-sectional anonymous phone surveys using Computer Assisted Telephone Interview (CATI) methodology, with a nationally representative sample of at least 1000 participants each month. The recorded population was representative in terms of gender, age, education level, and type of settlement. Sampling errors were further corrected by post-stratification weights. The data collection adhered to European and Hungarian privacy regulations, approved by the Hungarian National Authority for Data Protection and Freedom of Information [47], as well as the Health Science Council Scientific and Research Ethics Committee (resolution number IV/3073-1/2021/EKU) See SI Section 1.2. for further details).

## Data Availability

All data needed to evaluate the conclusions in the paper are present in the paper and/or the Supplementary Materials.

## Acknowledgments

M.K. and A.M. are grateful to Júlia Koltai and Gergely Röst for their contribution in collecting and sharing the MASZK dataset.

## Funding

A.M. and M.K. were supported by the Accelnet-Multinet NSF grant. A.M. is grateful for the support from CEU and NetSI (Northeastern University). M.K. acknowledges support from the ANR project DATAREDUX (ANR-19-CE46-0008); the SoBigData++ H2020-871042; the EMOMAP CIVICA projects; and the National Laboratory for Health Security, Alfréd Rényi Institute, RRF-2.3.1-21-2022-00006.

## Competing interests

The authors declare that they have no competing interests.

## Author’s contributions

A.M. performed the numerical simulations and data analysis. A.M. and N.P. developed the analytical formulation. A.M. wrote the first draft of the manuscript. All authors designed the study, interpreted the results, edited, and approved the manuscript.

## Supplementary Information

### 1 Data

#### 1.1 Vaccination data

Data on the number of vaccines administered have been sourced from Our World in Data [1]. Figures S1 presents the number of vaccines administered daily by age for Hungary.

**Figure S1:**
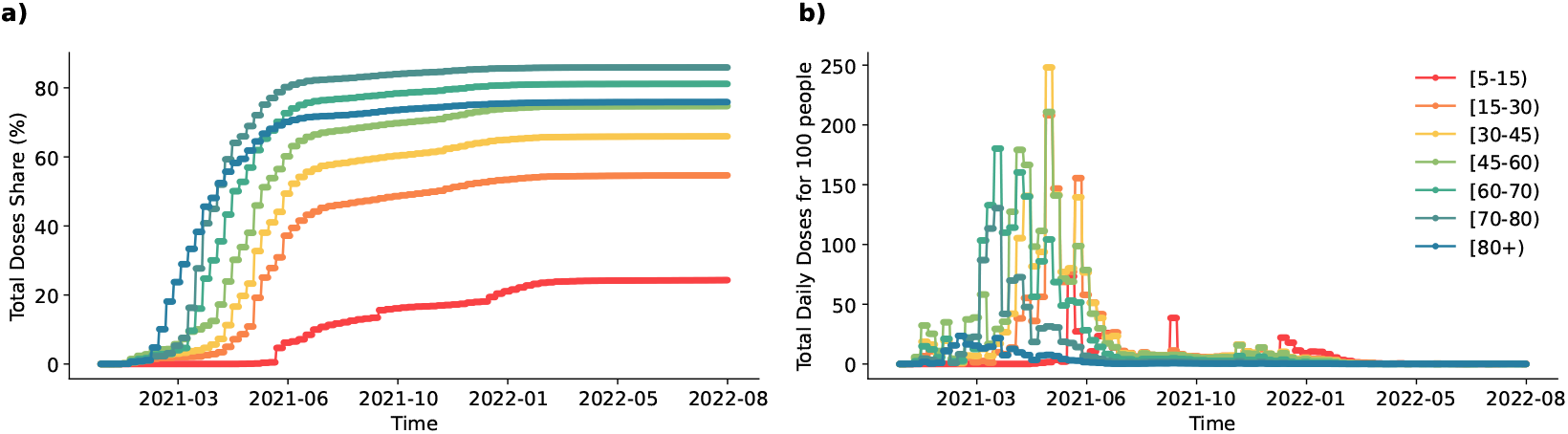
Hungary COVID-19 vaccination. **a.** Share of people who completed the initial COVID-19 vaccination protocol by age **b**. Total Daily Dosed for 100 people by age.

#### 1.2 Contact data

In this study, we used data coming from the MASZK survey study [2, 3], a large data collection effort on social mixing patterns made during the COVID-19 pandemic, conducted in Hungary from April 2020 to July 2022. The study involved 26 monthly cross-sectional anonymous phone surveys using Computer Assisted Telephone Interview (CATI) methodology, with a nationally representative sample of 1000 participants each month. The recorded population was representative in terms of gender, age, education level, and type of settlement. Sampling errors were further corrected by post-stratification weights. The data collection adhered to European and Hungarian privacy regulations, approved by the Hungarian National Authority for Data Protection and Freedom of Information [4], as well as the Health Science Council Scientific and Research Ethics Committee (resolution number IV/3073-1/2021/EKU).

Relevant to this study, the questionnaires recorded information about the *proxy social contacts*, defined as interactions where the respondent and a peer stayed within 2 meters for more than 15 minutes [5], at least one of them without wearing a mask. Approximate contact numbers were recorded between the respondents and their peers from different age groups of 0–4, 5–14, 15–29, 30–44, 45–59, 60–69, 70–79, and 80+. Contact number data about underage children were collected by asking legal guardians to estimate daily contact patterns. Participants during the whole data collection were asked to report contacts referring (*i*) to the previous day and, during the first data collection campaigns (*ii*) to an average pre-pandemic day (that we use for the analysis in Fig 1-4 in the main text). Additionally, in three data collection waves: April 2021, November 2021, and June 2022 contacts have been collected in the form of diaries (that we use for the analysis in Fig 5 in the main text). Namely, participants were asked to list one by one the contacts they had on the previous day by providing some socio-demographic information about the contacts such as their wealth situation. Beyond information on contacts before and during the pandemic, the MASZK dataset provided us with information on *social-demographic characteristics* of participants, such as their *perceived wealth situation, gender, vaccination status*, etc.

### 2 Epidemic models

This section provides a detailed description of the SEIR model with vaccination. Susceptible individuals, in contact with the infected, might be exposed to the virus with a rate driven by the force of infection Λ; exposed are not yet infectious and transition to the infected compartment with rate Ψ; infected individuals recover with rate Γ. Each compartment is divided between non-vaccinated *NV* and vaccinated *V*. Thus the total number of individuals in compartment *X* (i.e., *X* = [*S, E, I, R*]) is *X* = *X*_*NV*_ +*X*_*V*_. Individuals who received one dose of vaccine move to the compartments denoted with the superscript *V*. We assume all individuals except for the infectious can receive the vaccine. The number of people that get vaccinated at each time step is defined by Ω(*t*), and it is distributed among *S*_*NV*_, *E*_*NV*_, *R*_*NV*_, proportionally to their share in the population. The effectiveness of the vaccine is described by *g*_1_ and *g*_2_ which respectively represent efficacy against infection and death. Additionally, we assume that infected vaccinated individuals are less likely to further transmit the virus, this is modelled by the parameter *V E* in the force of infection. The formulation of this model varies depending on how the population structure and contact patterns are described. Below, we present two different models that we study.

**Figure S2:**
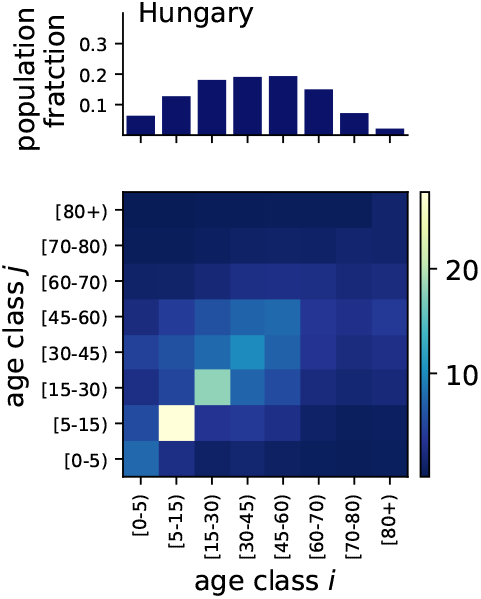
(1*st row*) Population distribution by age (2*nd row*) Age-contact matrices referred to the pre-pandemic period for Hungary.

#### 2.1 Age-stratification

Standard approaches to model the spreading of infectious diseases often acknowledge the stratification of contacts across age brackets. To this end, contact matrices **C** are introduced. The element *C*_*ij*_ quantifies the average number of contacts that an individual in age-bracket *i* has with individuals in age group *j* within a certain time window [6–8]. The population is divided into age brackets so that 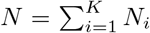. The variables *N*_*i*_ capture the number of individuals in age group *i* while *K* indicates the number of different age groups. The SEIR model with vaccination that incorporates such matrices can be written as follows:

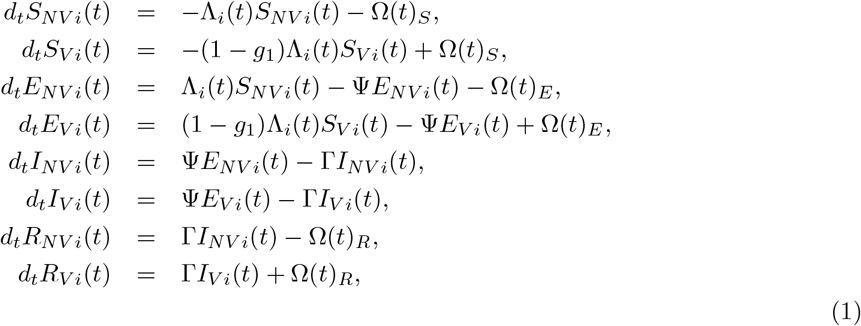

where *i* is the index that describes the membership of individuals in the *K* age groups and the subscripts *NV* and *V* indicate respectively the non-vaccinated and vaccinated individual in each compartment. The force of infection is then defined as the per-capita rate at which susceptibles acquire infections:

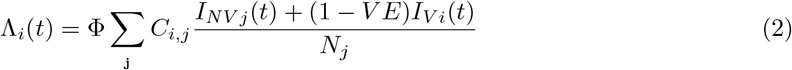

#### 2.2 Generalized models

The SEIR model with vaccination featuring generalized contact matrices [9] considers the population sliced in *m* +1 dimensions. Age, plus *m* others dimensions. Here, we have considered *m* = 1, hence two dimensions in total. The epidemic dynamics are encoded in the following set of differential equations:

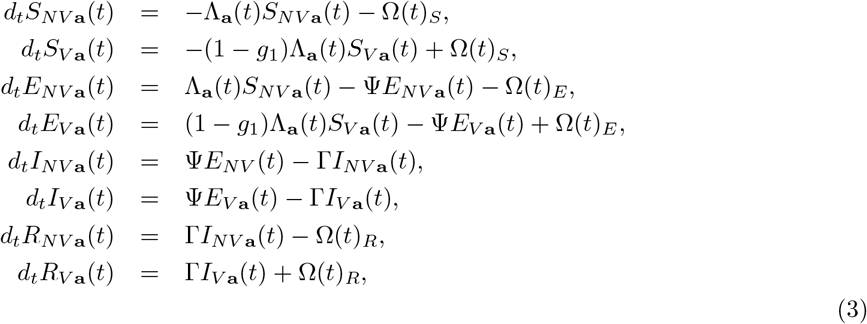

where **a** = (*i, α*, …, *γ*) is the index vector that describes the membership of individuals in the *m* + 1 groups and the subscripts *NV* and *V* indicate respectively the non-vaccinated and vaccinated individual in each compartment. The force of infection can be written as:

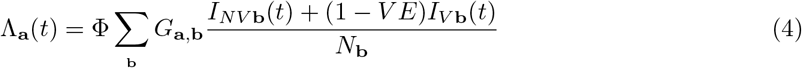

where Φ is the transmissibility of the disease, *V E* is the vaccine efficacy against further trasmission. The temporal dependence is induced by the variation in the number of infected across age brackets.

##### 2.2.1 Modeling vaccination uptake among the second dimension

As shown in Figure S1, vaccination data on the daily doses administered is available, at the lower level, by age groups. To introduce a bias in the vaccination uptake among individuals in the second dimension, we use true data stratified by age and distribute it along the second dimension according to a given distribution *P*_*vax*_(*α*), which indicates the proportion of vaccines allocated to each subgroup. Let Ω_*i*_(*t*) represent the daily administered doses at time *t* for age group *i*. The number of vaccines administered to a subgroup *α* is then Ω_*i,α*_(*t*) = Ω_*i*_(*t*) · *P*_*vax*_(*α*).

We assume that each subgroup can be vaccinated up to 95%. If this limit is reached, any excess vaccinations are redistributed randomly within that particular age group.

### 3 Computing attack rates

The attack rate is defined as the fraction of all individuals in a susceptible population who have been infected before a specific time *t*. In a SEIR model, the attack rate at time *t* can be computed as follows:

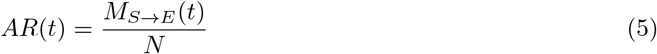

where *M*_(*S*→*E*)_ indicates the number of transitions from the susceptible (*S*) to the exposed (*E*) state up to time *t*, and *N* represents the total number of individuals in the population.

However, when vaccination is introduced, the computation of the attack rate for *vaccinated* and *non-vaccinated* individuals separately becomes more complex. In particular, when vaccination is administered during the course of the epidemic, not only are susceptible individuals vaccinated but also those who have been exposed (i.e., already infected but not yet infectious) and those who have recovered. In epidemiological modeling, this means that over the course of the epidemic, there are continuous movements between *S*_*NV*_ →*S*_*V*_ but also *E*_*NV*_ →*E*_*V*_ and *R*_*NV*_ →*R*_*V*_.

When computing the attack rate for *vaccinated* and *non-vaccinated* individuals separately, it is important to consider these transitions and account for the fact that vaccination is also administered to individuals in the *E*_*NV*_ and *R*_*NV*_ compartments. Therefore, the denominator of the attack rate is not simply the total number of vaccinated and non-vaccinated individuals at time *t*, but it should also account for these transitions.

#### Non-vaccinated population

For the non-vaccinated population, the denominator should include not only the total number of individuals not yet vaccinated at time *t* but also all the transitions from *E*_*NV*_ → *E*_*V*_ and *R*_*NV*_ → *R*_*V*_ up to time *t*. This is because these individuals, although vaccinated later, were not vaccinated when they were exposed or recovered. Thus, the adjusted denominator is:

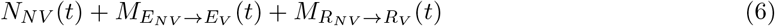

Where *N*_*NV*_ (*t*) is the total number of non-vaccinated individuals at time *t*, and 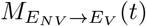 and 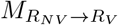 indicate the number of individuals vaccinated up to time *t* in the exposed and recovered compartments, respectively. As *N*_*NV*_ (*t*) can also be expressed as the difference between the total population and the vaccinated individuals, *N* − *N*_*V*_ (*t*), and *N*_*V*_ (*t*) can be expressed as the sum of all the vaccinated individuals, ie 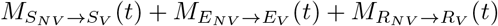, we can rewrite Equation (6) as the difference between the total population and the number of individuals vaccinated in the susceptible compartment, the attach rate can be expressed as follow.

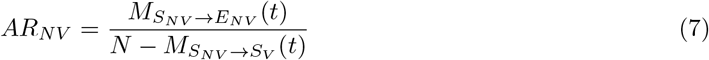

#### Vaccinated population

Symmetrically, the denominator of the attack rate for vaccinated individuals should account for the fact that individuals moving from *E*_*NV*_ to *E*_*V*_ and *R*_*NV*_ to *R*_*V*_ were not vaccinated when they were initially infected. Therefore, these individuals should be removed from the count of the total number of vaccinated individuals at time *t*. Thus, the denominator becomes:

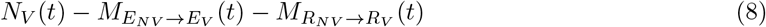

By substituting *N*_*V*_ (*t*) in Equation (8), we can compute the attach rate as follow:

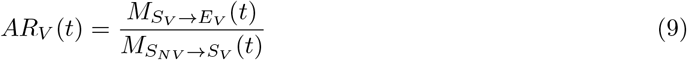

#### Entire population

Finally, the overall attack rate for the entire population can be computed as follows:

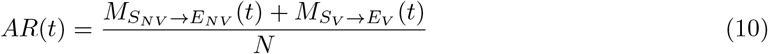

### 4 Inferring the burden of the epidemic for subgroups from age-stratified model output

To estimate the epidemic burden for specific subgroups *α* using outputs from a model stratified solely by age, we first need to infer the epidemic burden within each age group in the subgroup (*i, α*) and subsequently aggregate these age-specific results to obtain *AR*_*α*_. This requires estimating the number of susceptible individuals in each group defined by age and SES who became exposed during the epidemic i.e., 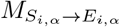.

Assuming knowledge of the number of vaccinated individuals across subgroups defined by age and SES, one can infer this number using age-stratified output of traditional models. Namely, this involves computing the attack rate for both vaccinated and unvaccinated individuals, as described in Equations (9) and (7) in each age group *i*, from the age-stratified model. Therefore, we can multiply *AR*_*NV i*_ and *AR*_*V i*_ by the total number of non-vaccinated and vaccinated individuals in each group (*i, α*) at time *t*. However, in line with the considerations explained in the previous section, it is important to account for the transitions *E*_*NV i*_ → *E*_*V i*_ and *R*_*NV i*_ → *R*_*V i*_ that occur over the course of the epidemic. The equation used for this calculation reads as follows:

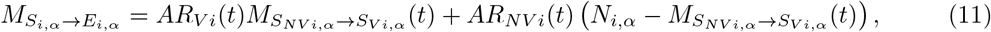

where *AR*_*V i*_(*t*) and *AR*_*NV i*_(*t*) represent respectively the attack rates for vaccinated and non-vaccinated individuals in age group *i* and 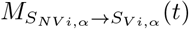 denotes the number of susceptible individuals in groups (*i, α*) who were vaccinated at time *t*. It is important to note that in a real-world case, we can estimate the number of vaccines administered to those who have already recovered from the infection, especially if they were symptomatic. However, it is not possible to determine the number of vaccines administered to individuals who were exposed. The attack rate for groups (*i, α*) can be then calculated by dividing equation (11) for the population size as follows:

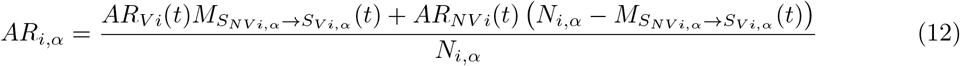

Finally, we can then aggregate by age groups equation (12) and divide by the total number of individuals in subgroup *α* as follows:

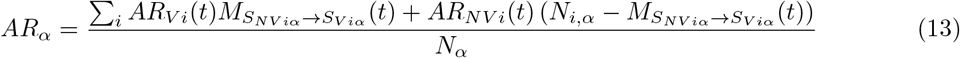

### 5 Synthetic population and mixing patterns

The age contact matrix and population distribution used are based on data for Hungary, as illustrated in Figure S2. The model used to build the synthetic population and their mixing pattern (Figures 1 − 4) is the same as in Ref. [9]. Here, we report the values of the activity, assortativity and the free parameters (see Table S1).

**Table S1:** List of parameters used to generate Figure 2 in the main text.

| SES Group | 1 | 2 | 3 |
| --- | --- | --- | --- |
| Population distribution | 35% | 45% | 20% |
| Assortativity | 60% | 50% | 65% |
| Activity | 20% | 40% | 40% |
| Free parameters | 0.6 | 0.6 | 0.5 |

### 6 Robustness analysis for Figures 1 and 3

In this section, we present a series of alternative simulation settings for Figures 1 and 3 of the main text to assess the robustness of the results. By exploring different assumptions and parameter variations, we aim to ensure that the findings remain consistent across a range of plausible scenarios. These robustness checks provide additional confidence in the conclusions drawn from the main simulations.

#### 6.1 Vaccination stratified only by SES

Here, we simulate vaccination distributions that do not account for age, by only applying the vaccination distributions *V D*1, *V D*2, and *V D*3 across three socioeconomic status (SES) groups. This approach is applied to both Scenario 1 and Scenario 2, as shown in Figures S3 and S4, respectively.

In these simulations, the variations among the three vaccination distributions (VDs) are more pronounced in terms of overall prevalence across the population, as observed in panel *a* of Figures S3 and S4. Notably, in Scenario 2, the effectiveness of *V D*2 and *V D*3 increases substantially, nearly eliminating the epidemic wave.

Furthermore, differences among SES subgroups’ attach rate become even more evident, as shown in panel *b* of Figures S3 and S4.

Finally, panels *c* of Figures S3 and S4 demonstrate that attack rates for specific subgroups are not accurately predicted by the age-stratified model. Indeed, the discrepancy between the two methodologies, *G*_**ab**_ − *f* (*C*_*ij*_), is almost always non-zero.

Overall, these simulation results align with the findings discussed in the main text, supporting the conclusions on the influence of vaccination distribution patterns on epidemic outcomes.

**Figure S3:**
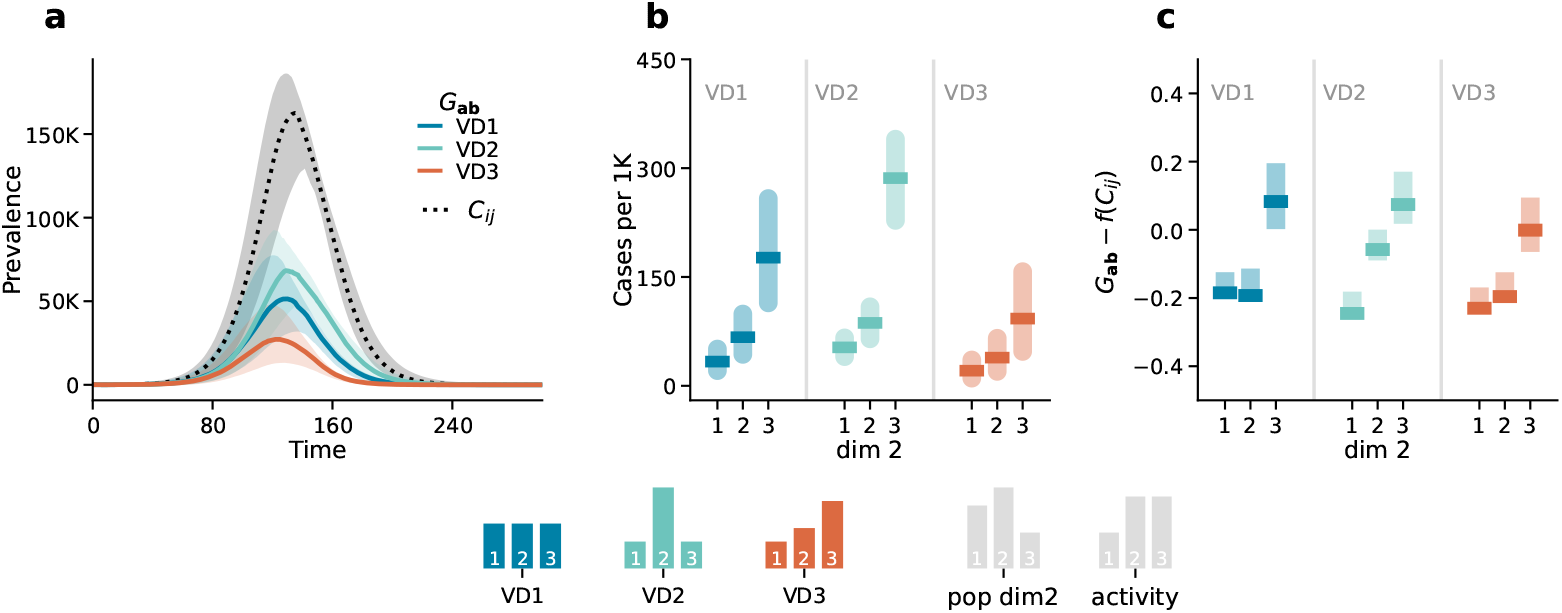
Scenario 1. Epidemic outcomes and predictability: Panel *a* displays the number of newly infected individuals over time. The dotted black line corresponds to the outcome of the age-stratified model (*C*_*ij*_), and the solid coloured lines correspond to the outcome of the generalized model (*G*_**ab**_) in the three different vaccination strategies (*V D*1, *V D*2, *V D*3) Panel *b* shows the attack rate by the second dimension (dim 2) predicted by the generalized model (*G*_**ab**_. Panel *c* shows the difference between the attack rate predicted by the generalized model *G*_**ab**_ and the one estimated from the aggregate output of the age-stratified model *f* (*C*_*ij*_ by the second dimension (dim 2). Results refer to the median of 500 runs with IQRs. Epidemiological parameters: Γ = 0.25, Ψ = 0.4, *g*_1_ = 0.6, *R*_0_ = 1.6. Simulations start with *I*_0_ = 100 initial infectious seeds.

**Figure S4:**
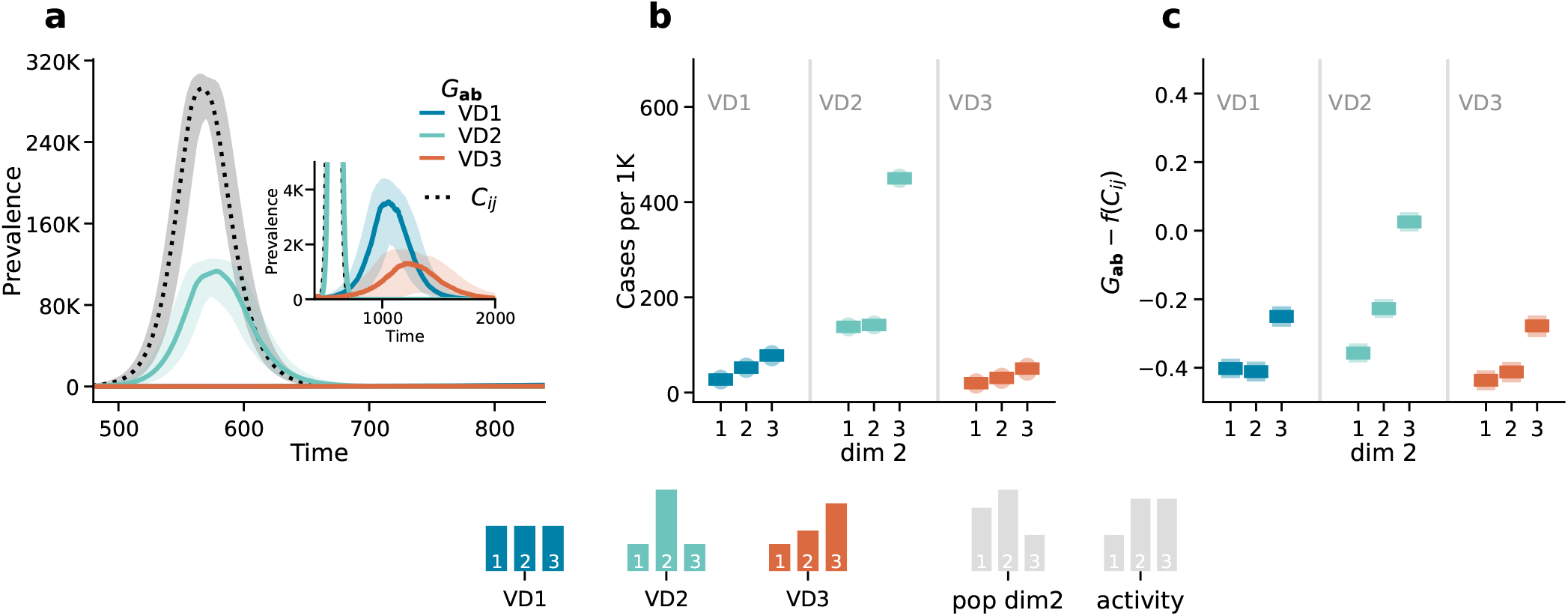
Scenario 2. Epidemic outcomes and predictability: Panel *a* displays the number of newly infected individuals over time. The dotted black line corresponds to the outcome of the age-stratified model (*C*_*ij*_), and the solid coloured lines correspond to the outcome of the generalized model (*G*_**ab**_) in the three different vaccination strategies (*V D*1, *V D*2, *V D*3) Panel *b* shows the attack rate by the second dimension (dim 2) predicted by the generalized model (*G*_**ab**_. Panel *c* shows the difference between the attack rate predicted by the generalized model *G*_**ab**_ and the one estimated from the aggregate output of the age-stratified model *f* (*C*_*ij*_ by the second dimension (dim 2). Results refer to the median of 500 runs with IQRs. Epidemiological parameters: Γ = 0.25, Ψ = 0.4, *g*_1_ = 0.6, *R*_0_ = 3. Simulations start with *I*_0_ = 100 initial infectious seeds.

##### 6.1.1 Homogeneous mixing

Keeping the same vaccination distributions, we run the same analysis as present in Figure 1 (Scenario 1) and Figure 3 (Scenario 2) of the main text on a simpler population mixing setting. Furthermore, we consider vaccination only by SES as in the previous section. In this case, we assume homogeneous (i.e., random) mixing where contacts along the second dimension are set proportional to the product of the population sizes in each group. The only set of parameters of interest is the population distribution, which is set to 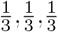. Indeed, in these settings, no extra parameters are needed. Again the model is the same as in Ref. [9].

###### Scenario 1

The first observation is that, as expected from Ref. [9], homogeneous mixing leads to the same epidemic dynamics (i.e., same prevalence) across subgroups (see panels S5*a*). Indeed, regardless of the vaccination distribution the coloured lines overlap. Furthermore, random mixing leads to the same total prevalence regardless of the inclusion of a second dimension in the contact matrices. Indeed, solid coloured and dashed grey lines representing respectively the prevalence estimated by modes fed with generalised or traditional contact matrices overlap.

In panels *b* of Figure S5 we show the attack rate per one thousand (i.e., number of cases for 1000 individuals) for each subgroup, for each vaccination distribution (VDs). In a random mixing situation, differences in disease burden among subgroups arise solely from variations in vaccination uptake. When vaccination uptake is equal across all groups (*V D*1), infection rates are the same in all three SES groups. Under vaccination distribution *V D*2, the second SES group shows the lowest infection rate due to higher vaccination uptake. In the more unequal distribution of vaccines (*V D*3), the number of cases in each group inversely correlates with their respective vaccination uptake: the first group, having the lowest vaccination rate, becomes the most infected, while the third group, with the highest vaccination uptake, experiences the fewest cases.

Finally, in panel S5*c* we show the difference of the number of cases in each subgroup from the generalized model *G*_**ab**_ and as estimated from a simple age-stratified model *f* (*C*_*ij*_). In this scenario, the differences among the two techniques, although small, are almost always different from zero.

**Figure S5:**
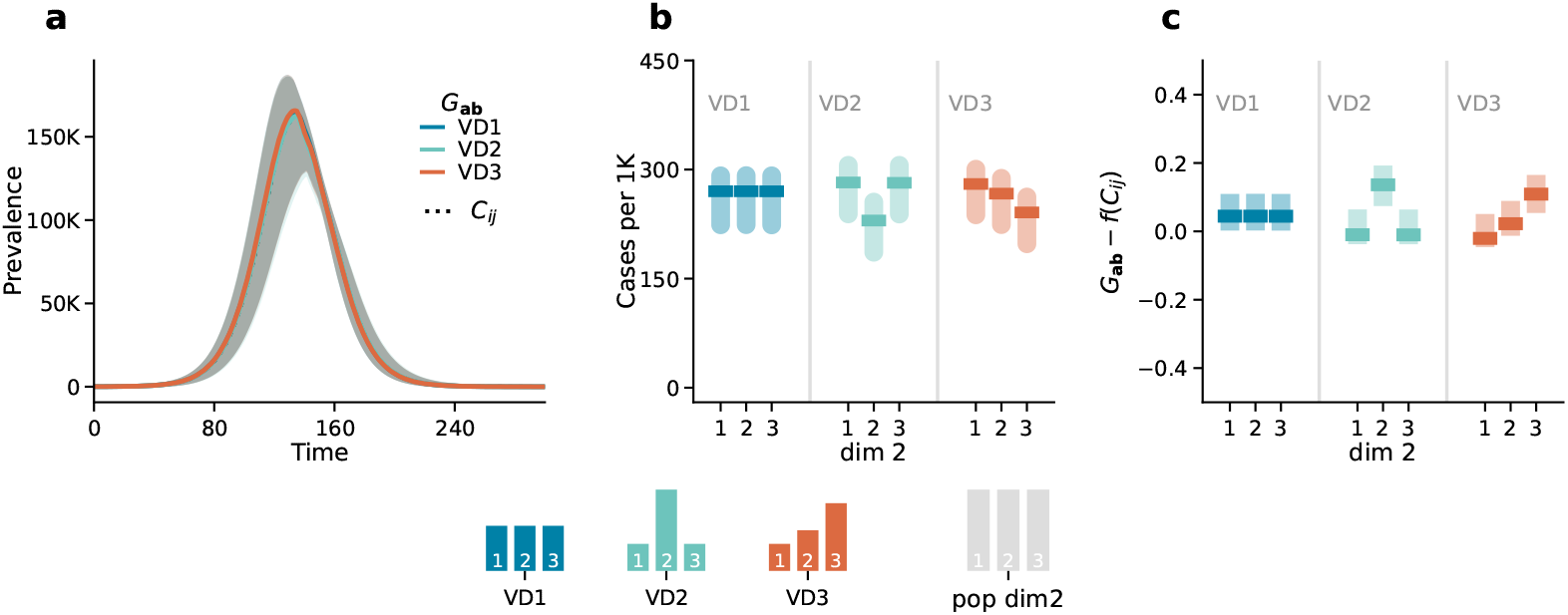
Scenario 1. Epidemic outcomes and predictability: Panel *a* displays the number of newly infected individuals over time. The dotted black line corresponds to the outcome of the age-stratified model (*C*_*ij*_), and the solid coloured lines correspond to the outcome of the generalized model (*G*_**ab**_) in the three different vaccination strategies (*V D*1, *V D*2, *V D*3) Panel *b* shows the attack rate by the second dimension (dim 2) predicted by the generalized model (*G*_**ab**_. Panel *c* shows the difference between the attack rate predicted by the generalized model *G*_**ab**_ and the one estimated from the aggregate output of the age-stratified model *f* (*C*_*ij*_ by the second dimension (dim 2). Results refer to the median of 500 runs with IQRs. Epidemiological parameters: Γ = 0.25, Ψ = 0.4, *g*_1_ = 0.6, *R*_0_ = 1.6. Simulations start with *I*_0_ = 100 initial infectious seeds.

###### Scenario 2

Here, we report the results of the same simulation setting for Scenario 2, where the vaccination campaign rollout happened before the epidemic outbreak.

Panel *a* of Fig. S6 shows that similar to Scenario 1, the coloured lines overlap regardless of the vaccination distribution, indicating consistent epidemic outcomes across different distribution strategies. Additionally, these coloured lines match the dashed grey lines, which represent outcomes based on traditional age contact matrices, highlighting that the introduction of an additional dimension does not alter the overall results, as also discussed in [9].

Panel S6*b* shows the attack rate per one thousand (i.e., number of cases for 1000 individuals) for each subgroup, for each vaccination distribution (VDs). As before, in a random mixing situation, differences in disease burden among subgroups arise solely from variations in vaccination uptake. Thus the qualitative differences among attach rates in subgroups follow the ones seen in Scenario 1.

Panel S6*c* shows the difference of the number of cases in each subgroup from the generalized model *G*_**ab**_ and as estimated from a simple age-stratified model *f* (*C*_*ij*_). Interestingly, in this case, the difference is always zero, meaning that, in this case, it is possible to accurately predict the epidemic burden in the three additional subgroups starting from the output from age-stratified models. Indeed, in this scenario, since the vaccination is already completed before the epidemic begins, there is not a dynamic interaction between the vaccination uptake and the epidemic spreading, thus the attack rate in the different subgroups is fully captured by the overall number of individuals in each subgroup, that were vaccinated before the epidemic started.

**Figure S6:**
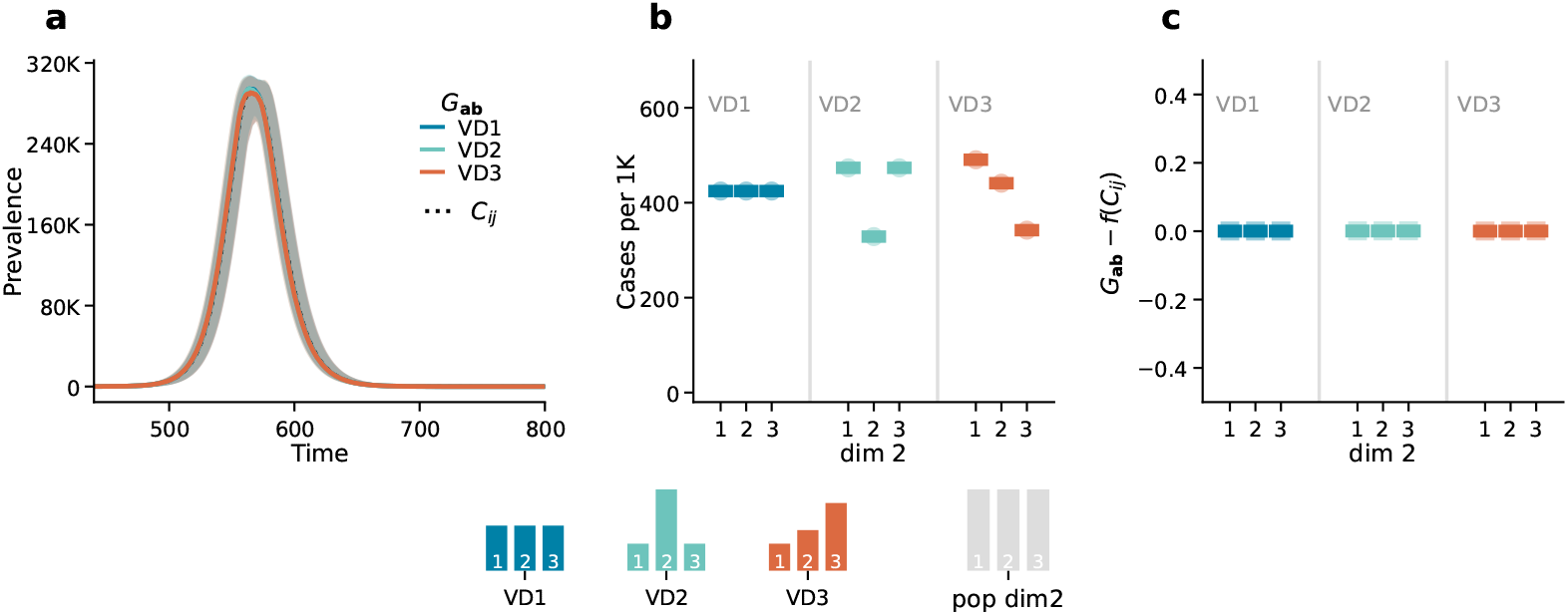
Scenario 2. Epidemic outcomes and predictability: Panel *a* displays the number of newly infected individuals over time. The dotted black line corresponds to the outcome of the age-stratified model (*C*_*ij*_), and the solid coloured lines correspond to the outcome of the generalized model (*G*_**ab**_) in the three different vaccination strategies (*V D*1, *V D*2, *V D*3) Panel *b* shows the attack rate by the second dimension (dim 2) predicted by the generalized model (*G*_**ab**_. Panel *c* shows the difference between the attack rate predicted by the generalized model *G*_**ab**_ and the one estimated from the aggregate output of the age-stratified model *f* (*C*_*ij*_ by the second dimension (dim 2). Results refer to the median of 500 runs with IQRs. Epidemiological parameters: Γ = 0.25, Ψ = 0.4, *g*_1_ = 0.6, *R*_0_ = 3. Simulations start with *I*_0_ = 100 initial infectious seeds.

##### 6.1.2 Sensitivity to varying *t*_*epi*_ and *R*_0_

Finally, we run a sensitivity analysis for the difference between the predicted number of cases in each subgroup from the generalized model and as estimated from a simple age-stratified model (*G*_**ab**_ − *f* (*C*_*ij*_)). Specifically, we examine the differences between the overall attack rate for each subgroup as predicted by the generalized model *G*_**ab**_ and the attack rate predicted by the outputs of the age-stratified model aggregate at age level, as described in Equation (13) *f* (*C*_*ij*_). In Figure S7, we show how this difference changes (*i*) as a function of *t*_*epi*_, the time at which the epidemic starts given that vaccination began at *t* = 0 with *R*_0_ = 2.5, and (*ii*) as a function of *R*_0_, the reproduction number, with *t*_*epi*_ = 0. Again the vaccination is kept to be stratified only by SES. These analyses are done both under a homogeneous mixing setting (panel *a*) and an assortative mixing setting (panel *b*). Both the parameters *t*_*epi*_ and *R*_0_ determine the overlap between the vaccination campaign and the epidemiological wave: with high *t*_*epi*_, vaccination might be completed before the epidemic starts, whereas with high *R*_0_, the epidemic might peak early during the vaccination campaign, diminishing the vaccination’s impact on the epidemic curve. The results indicate that only under random mixing, where vaccination is completed before the epidemic begins, can we accurately predict the attack rate within each subgroup based on the output of an age-stratified model (panel *a*, first column). Specifically, the difference (*G*_**ab**_ − *f* (*C*_*ij*_)) converges to zero as *t*_*epi*_ increases, indicating that in a fully vaccinated population with no interaction between epidemic and vaccination dynamics, the attack rate by subgroup aligns with predictions from the age-stratified model. However, as the overlap between the vaccination campaign and the epidemic increases, so does the error, as shown in panel *a*, first column, for low values of *t*_*epi*_ and in the second column for higher values of *R*_0_. In contrast, under assortative mixing, the interaction between differences in individual activity levels and vaccination uptake prevents accurate predictions (panel *b*).

**Figure S7:**
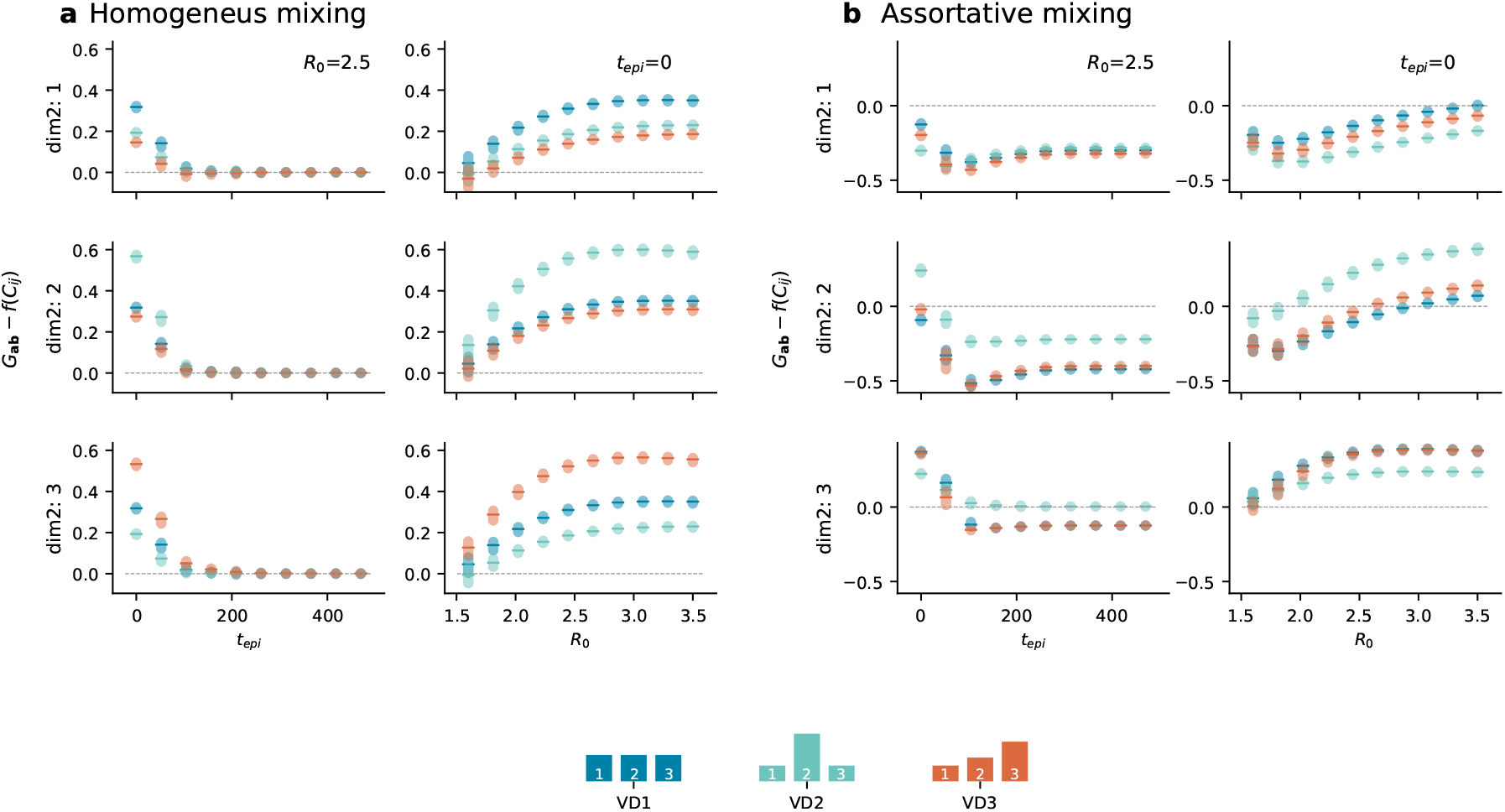
Sensitivity analysis to varying *t*_*epi*_ and *R*_0_. Differences between the overall attack rate for each subgroup as predicted by the generalized model *G*_*ab*_ and the attack rate predicted by the aggregate output of the age-stratified model, *f* (*C*_*ij*_) as a function of (i) *t*_*epi*_ with *R*_0_ = 2.5, and (ii) *R*_0_ with *t*_*epi*_ = 0. These analyses are done both under a random mixing setting (panel *a*) and an assortative mixing setting (panel *b*)

#### 6.2 Vaccination stratified by age and SES

Here, we explore a simulation setting with homogenous mixing while maintaining the vaccination modelling framework presented in the main analysis. Specifically, the distribution of vaccines follows a two-step approach: first by age, based on daily administration data, and then by socioeconomic status (SES), using three distinct vaccination uptake distributions (*V D*1, *V D*2, *V D*3).

##### 6.2.1 Homogeneous mixing

We run the same analysis as presented in Figure 1 (Scenario 1) and Figure 3 (Scenario 2) of the main text on a simpler population mixing setting. As before, in the case of homogeneous mixing contacts along the second dimension are set proportional to the product of the population sizes in each group. Again the model is the same as in [9].

The results for Scenario 1 and Scenario 2 are shown in Figure S8 and Figure S9, respectively. These findings are consistent with the patterns discussed in the previous section, further validating the dynamics observed when varying vaccination strategy and mixing assumptions.

**Figure S8:**
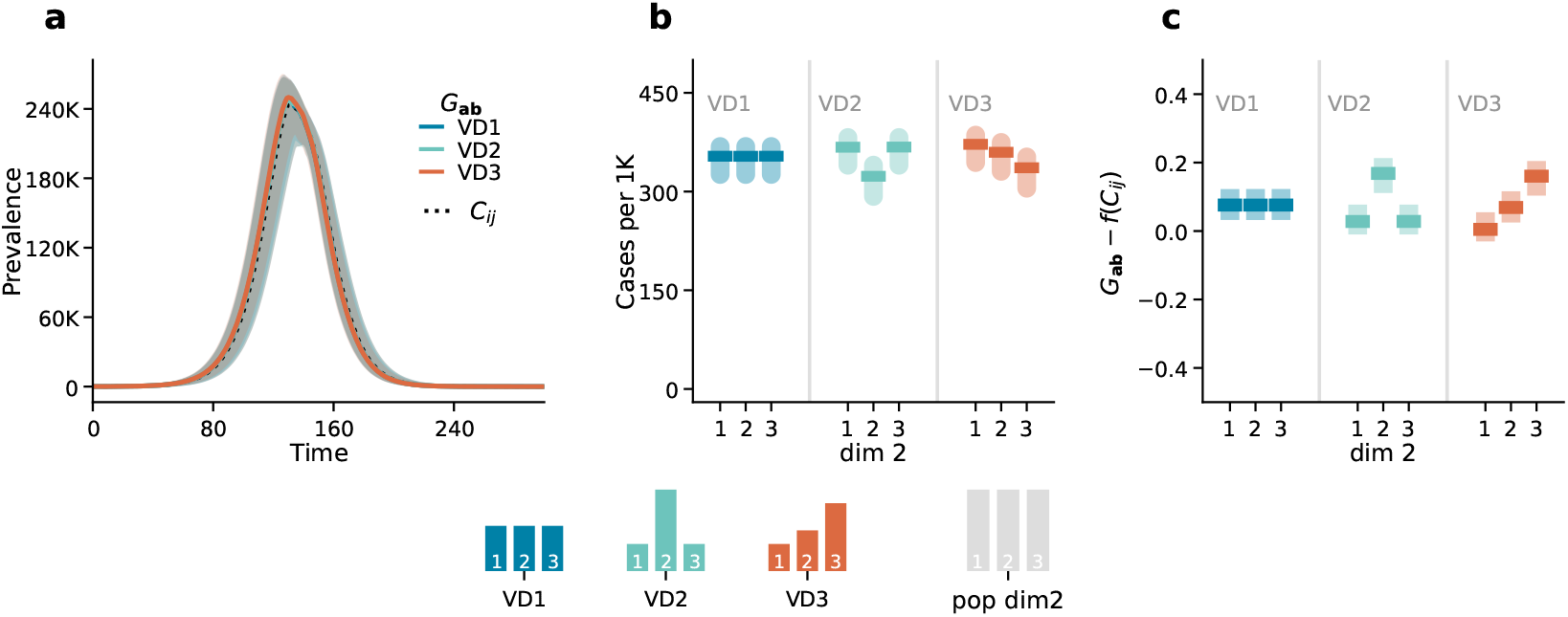
Scenario 1. Epidemic outcomes and predictability: Panel *a* displays the number of newly infected individuals over time. The dotted black line corresponds to the outcome of the age-stratified model (*C*_*ij*_), and the solid coloured lines correspond to the outcome of the generalized model (*G*_**ab**_) in the three different vaccination strategies (*V D*1, *V D*2, *V D*3) Panel *b* shows the attack rate by the second dimension (dim 2) predicted by the generalized model (*G*_**ab**_. Panel *c* shows the difference between the attack rate predicted by the generalized model *G*_**ab**_ and the one estimated from the aggregate output of the age-stratified model *f* (*C*_*ij*_ by the second dimension (dim 2). Results refer to the median of 500 runs with IQRs. Epidemiological parameters: Γ = 0.25, Ψ = 0.4, *g*_1_ = 0.6, *R*_0_ = 1.6. Simulations start with *I*_0_ = 100 initial infectious seeds.

**Figure S9:**
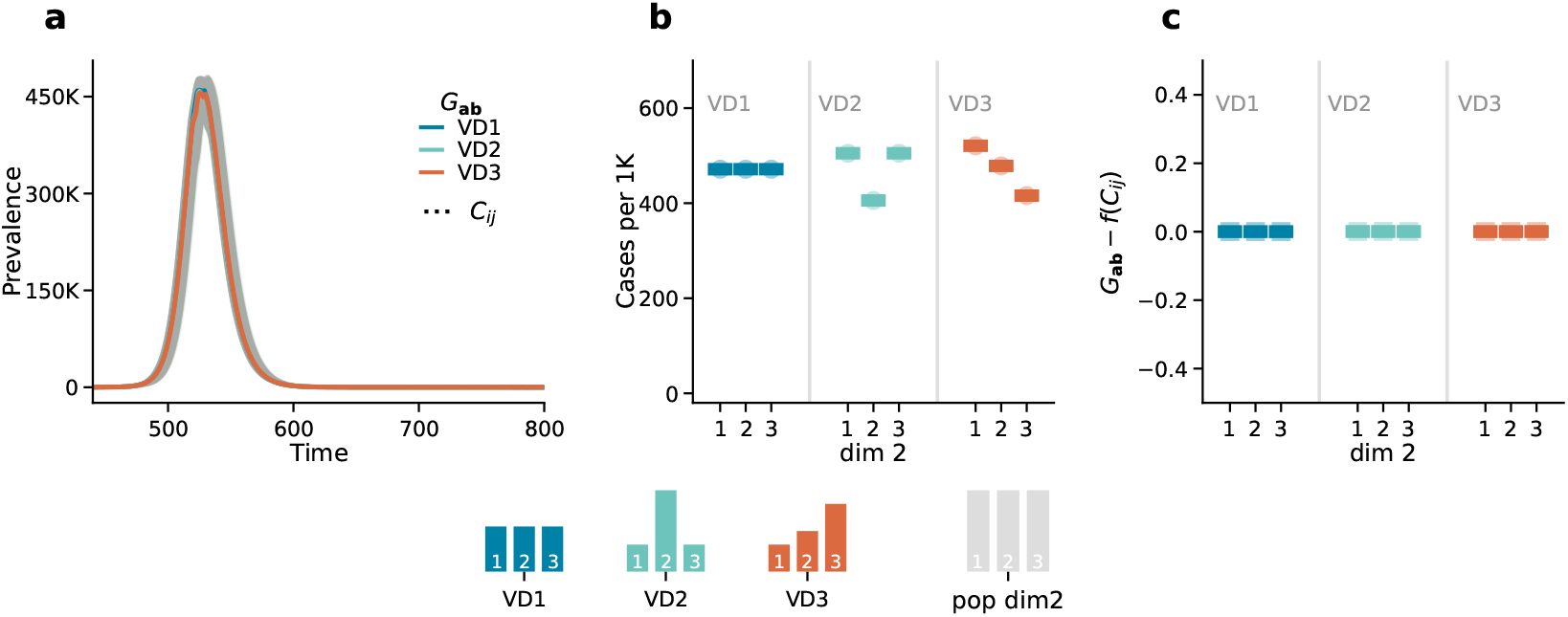
Scenario 2. Epidemic outcomes and predictability: Panel *a* displays the number of newly infected individuals over time. The dotted black line corresponds to the outcome of the age-stratified model (*C*_*ij*_), and the solid coloured lines correspond to the outcome of the generalized model (*G*_**ab**_) in the three different vaccination strategies (*V D*1, *V D*2, *V D*3) Panel *b* shows the attack rate by the second dimension (dim 2) predicted by the generalized model (*G*_**ab**_. Panel *c* shows the difference between the attack rate predicted by the generalized model *G*_**ab**_ and the one estimated from the aggregate output of the age-stratified model *f* (*C*_*ij*_ by the second dimension (dim 2). Results refer to the median of 500 runs with IQRs. Epidemiological parameters: Γ = 0.25, Ψ = 0.4, *g*_1_ = 0.6, *R*_0_ = 1.6. Simulations start with *I*_0_ = 100 initial infectious seeds.

##### 6.2.2 Sensitivity to varying *t*_*epi*_ and *R*_0_

We conducted the same sensitivity analysis for the difference between the predicted number of cases in each subgroup from the generalized model and as estimated from a simple age-stratified model (*G*_**ab**_ − *f* (*C*_*ij*_)), with results shown in Figure S10. As before, we examine both a homogeneous mixing setting (panel *a*) and an assortative mixing setting (panel *b*).

Consistent patterns emerge, leading to the same qualitative results and outcomes. Specifically, the results reaffirm that under random mixing, predictions of subgroup attack rates align closely with those from an age-stratified model when vaccination concludes prior to the epidemic onset. However, as the overlap between the vaccination campaign and the epidemic grows, discrepancies increase. This effect is particularly pronounced under assortative mixing, where the interaction between subgroup activity differences and vaccination uptake prevents accurate predictions.

**Figure S10:**
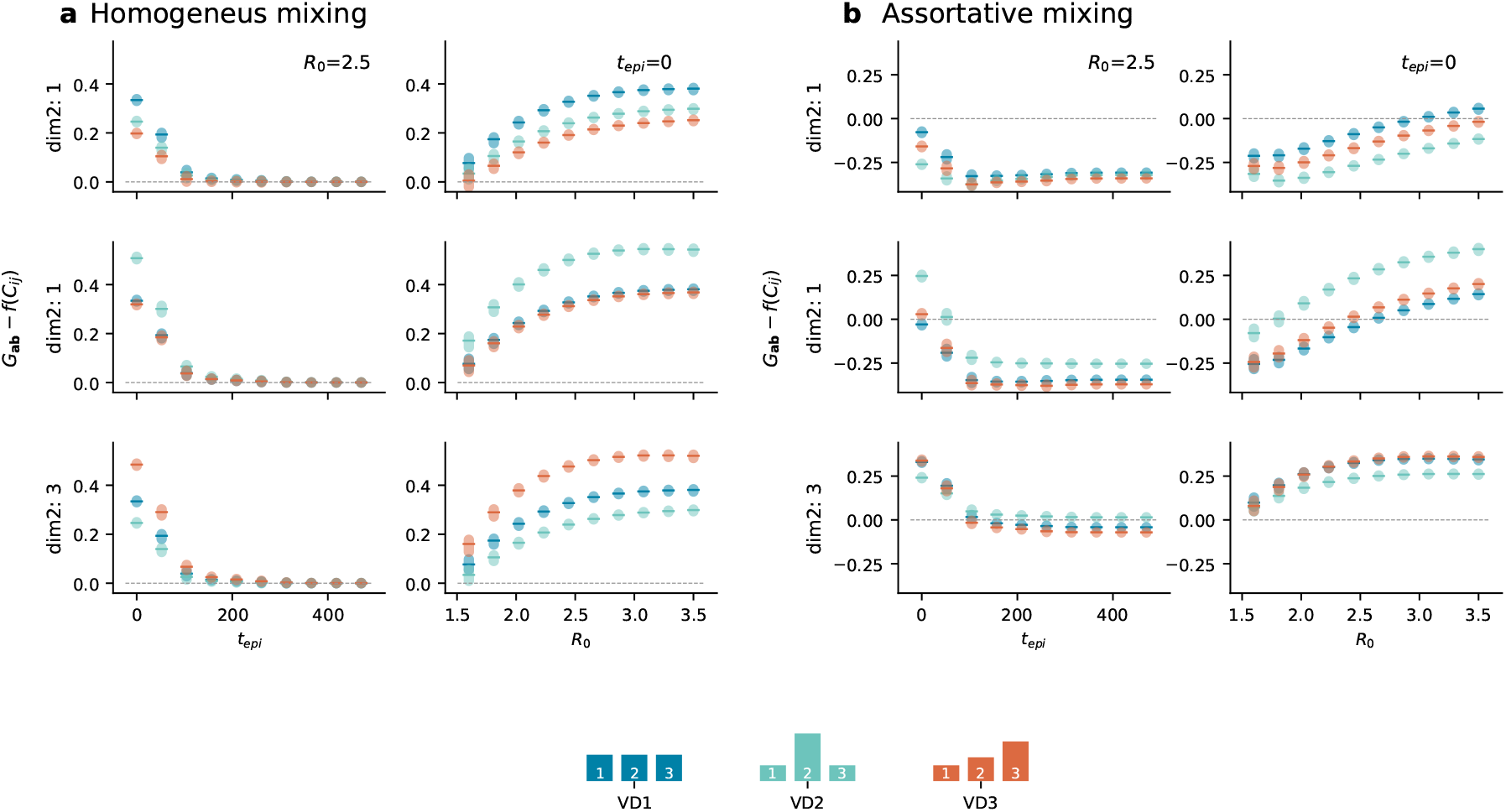
Sensitivity analysis. Differences between the overall attack rate for each subgroup as predicted by the generalized model *G*_*ab*_ and the attack rate predicted by the aggregate output of the age-stratified model, *f* (*C*_*ij*_) as a function of (i) *t*_*epi*_ with *R*_0_ = 2.5, and (ii) *R*_0_ with *t*_*epi*_ = 0. These analysis are done both under a random mixing setting (panel *a*) and an assortative mixing setting (panel *b*)

### 7 Non pharmaceutical interventions

Table S2 reports the values for activity, assortativity, and other free parameters used for both the tightened NPI simulation presented in Figure 2 (Scenario 1) and the relaxed NPI simulation presented in Figure 4 (Scenario 2) of the main text.

**Table S2:** List of parameters used to simulate the NPIs

| SES Group | 1 | 2 | 3 |
| --- | --- | --- | --- |
| Assortativity | 50% | 60% | 70% |
| Activity | 40% | 25% | 35% |
| Free parameters | 0.5 | 0.4 | 0.4 |

For completeness, we extend the model by adding a new compartment (*D*) to estimate the number of deaths under each vaccination distribution. In particular, we simulate the number of daily deaths by applying the age-stratified Infection Fatality Rate (*IFR*_*i*_) estimated for COVID-19 by Ref.[10]. The set of the equations presented in (3) can be extended as follows:

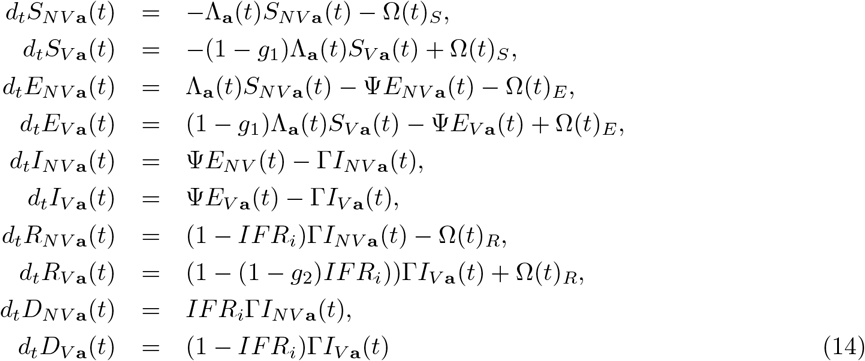

#### Section 1

We report the attack rate by socioeconomic status (SES) for both the Baseline setting and the setting with non-pharmaceutical interventions (NPI) in Figure S11.

**Figure S11:**
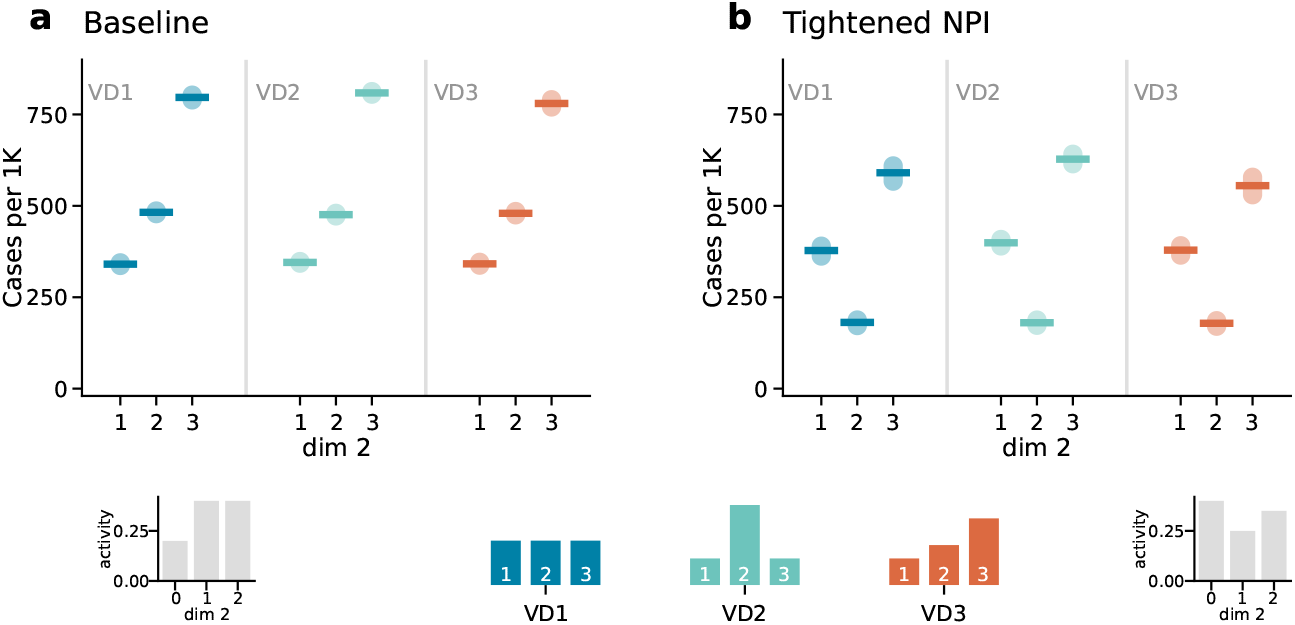
Scenario 1: The impact of NPIs on SES groups under different vaccination distributions. Panel *a* refers to the baseline case and shows the attack rate scaled by 1000 for the three SES. Panel *b* follows the same structure but refers to the case where tightened NPIs are introduced, reducing contacts by 20%. NPIs are adjusted 65 days after the epidemic onset. Results represent the median of 500 runs with confidence intervals. Epidemiological parameters: Γ = 0.25, Ψ = 0.4, *g*_1_ = 0.5, *g*_2_ = 0.8, and *R*_0_ = 2.5. Simulations begin with *I*_0_ = 100 initial infected seeds.

Figure S12 presents the results of the model with deaths for Scenario 1. In particular, panels *a* and *b* show respectively the prevalence in the baseline case and the case with tightened NPIs where contacts are reduced by 20%. Similarly, panels *c* and *d* display the total number of deaths in the baseline case and under tightened NPIs, respectively. The mortality trend exhibits the same qualitative behaviour as the prevalence.

**Figure S12:**
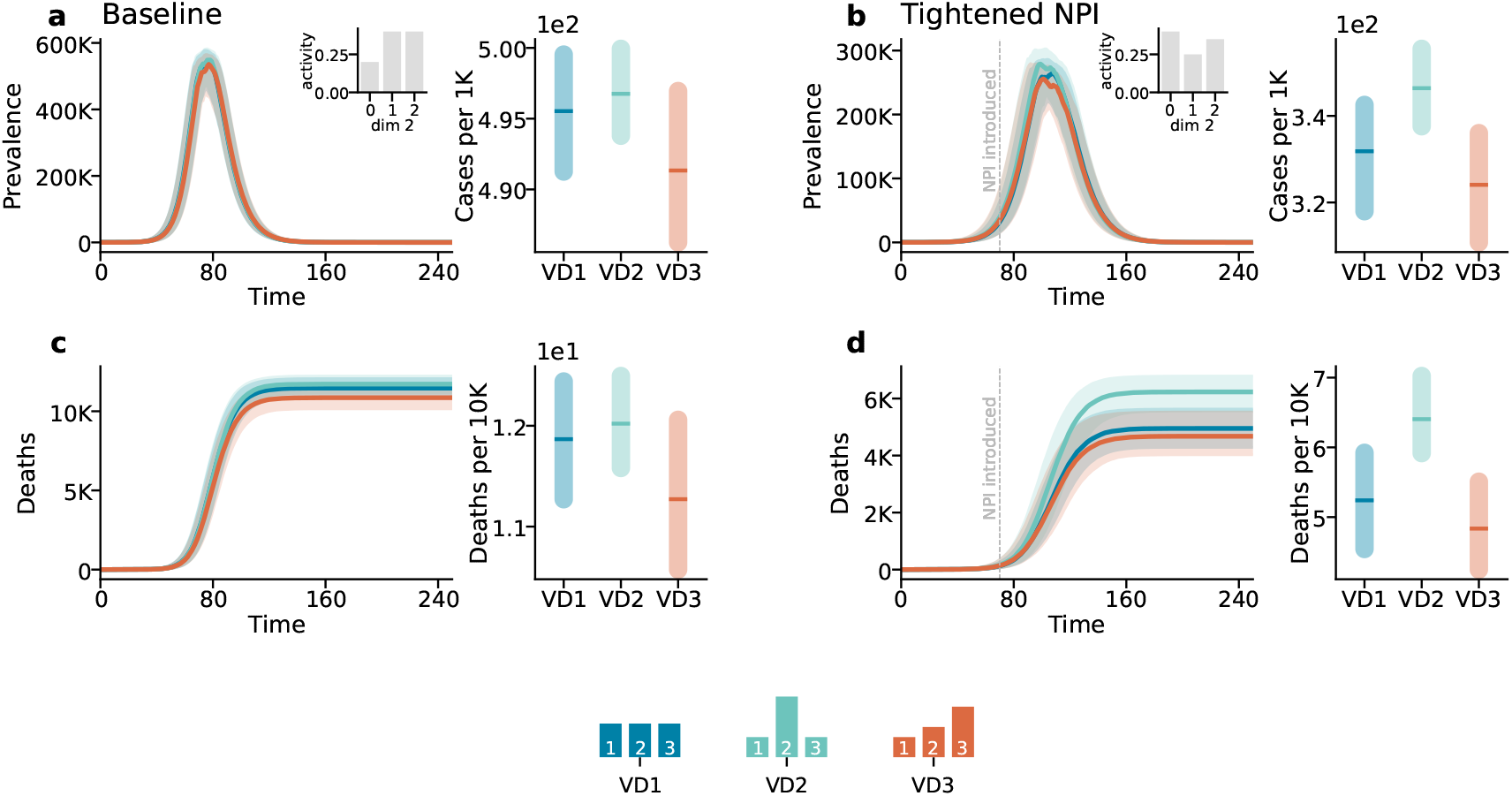
Scenario 1: The impact of NPIs under different vaccination distributions. Panel *a* refers to the baseline case, with insets depicting the activity distribution along the second dimension. The first column shows the total number of infected individuals, while the second column shows the attack rate scaled by 1000. Panel *b* follows the same structure but refers to the case where tightened NPIs are introduced, reducing contacts by 20%. Panels *c* and *d* follow the same structure but refer to the total number of deaths (first column) and mortality rate scaled by 10*K* (second column). NPIs are adjusted 65 days after the epidemic onset. Results represent the median of 500 runs with confidence intervals. Epidemiological parameters: Γ = 0.25, Ψ = 0.4, *g*_1_ = 0.5, *g*_2_ = 0.8, and *R*_0_ = 2.5. Simulations begin with *I*_0_ = 100 initial infected seeds.

#### Scenario 2

Similarly, we report the attack rate by socioeconomic status (SES) for both the Baseline setting and the setting with non-pharmaceutical interventions (NPI) in Figure S13.

While Figure S14 presents the results of the model that includes mortality for Scenario 2 following the same structure of Figure S12. Also in this case the mortality trend exhibits the same qualitative behavior as the prevalence.

**Figure S13:**
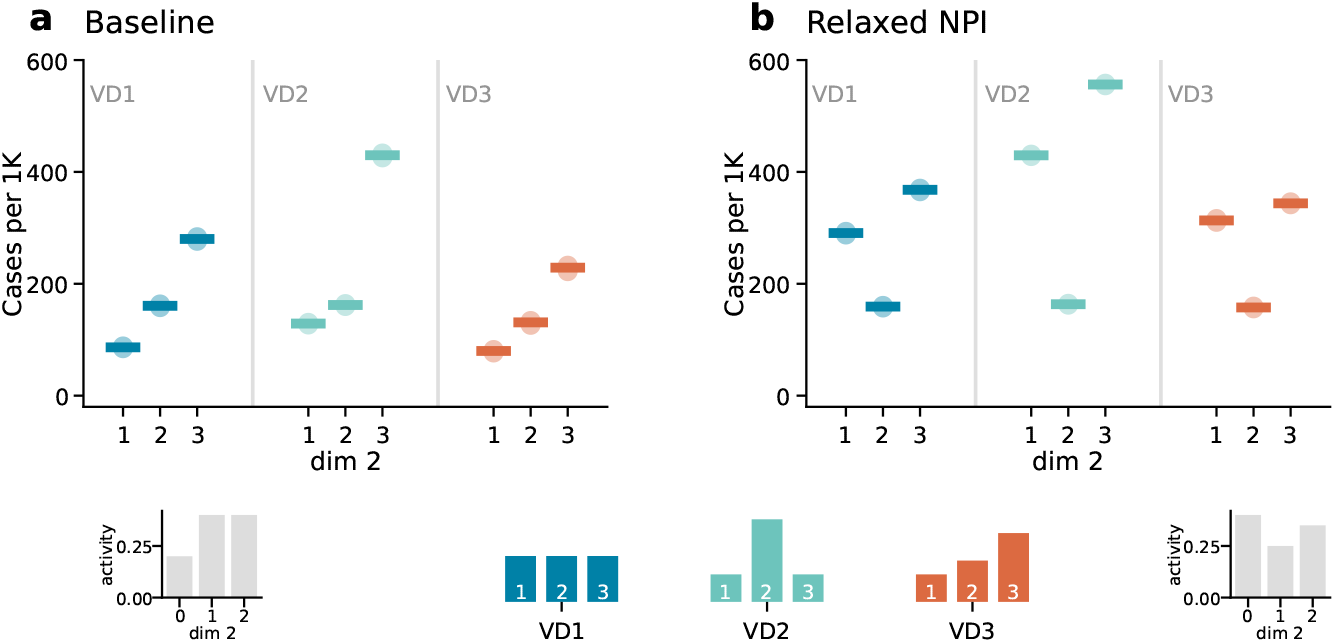
Scenario 2: The impact of NPIs on SES groups under different vaccination distributions. Panel *a* refers to the baseline case and shows the attack rate scaled by 1000 for the three SES. Panel *b* follows the same structure but refers to the case where tightened NPIs are released and the contacts increase by 20%. Panels *c* and *d* follow the same structure but refer to the total number of deaths (first column) and mortality rate scaled by 10*K* (second column). NPIs are adjusted 65 days after the epidemic onset. Results represent the median of 500 runs with confidence intervals. Epidemiological parameters: Γ = 0.25, Ψ = 0.4, *g*_1_ = 0.5, *g*_2_ = 0.8, and *R*_0_ = 2.5. Simulations begin with *I*_0_ = 100 initial infected seeds.

**Figure S14:**
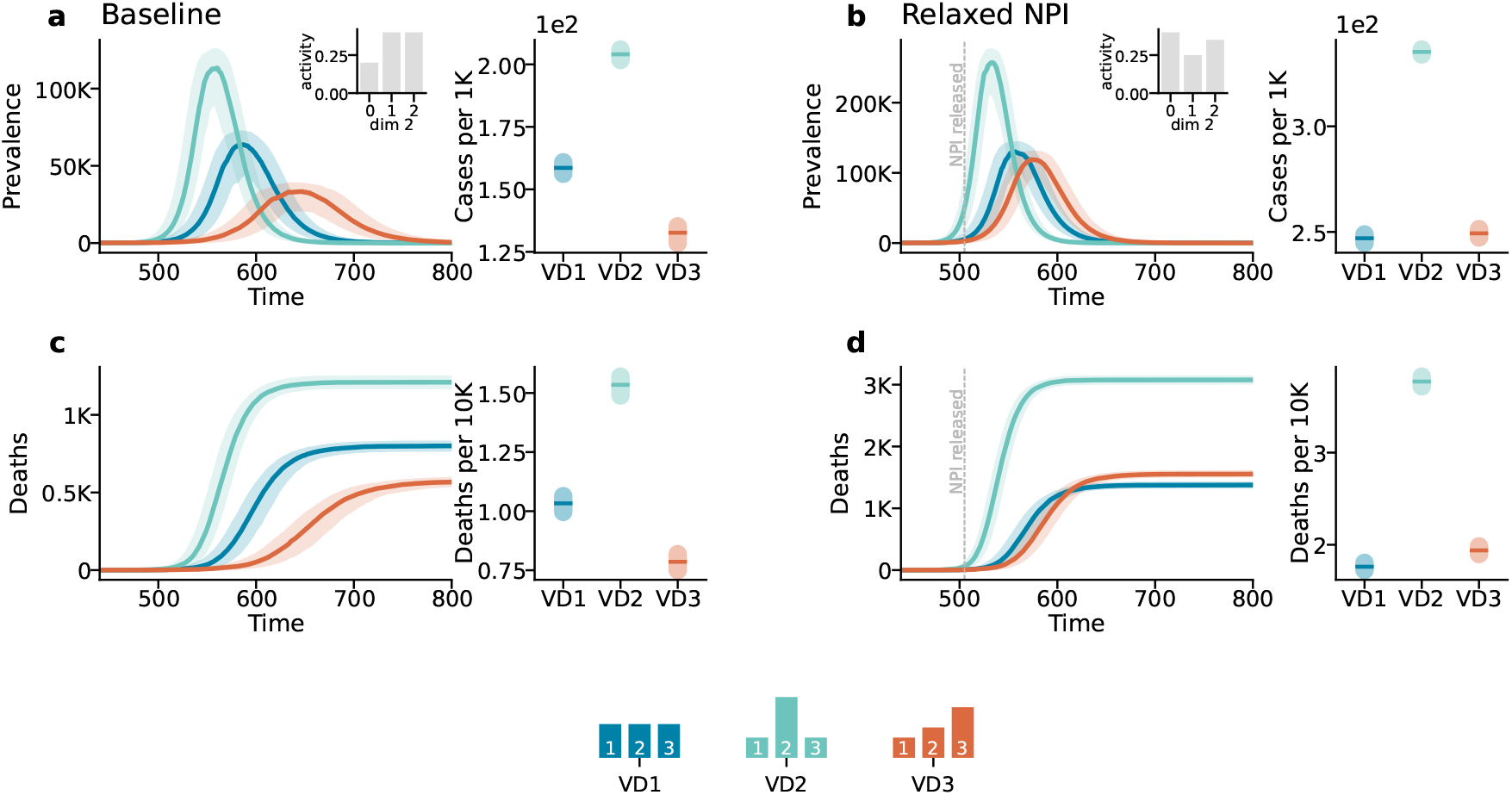
Scenario 2: The impact of NPIs under different vaccination distributions. Panel *a* refers to the baseline case, with insets depicting the activity distribution along the second dimension. The first column shows the total number of infected individuals, while the second column shows the attack rate scaled by 1000. Panel *b* follows the same structure but refers to the case where tightened NPIs are released, increasing contacts by 20%. NPIs are adjusted 65 days after the epidemic onset. Results represent the median of 500 runs with confidence intervals. Epidemiological parameters: Γ = 0.25, Ψ = 0.4, *g*_1_ = 0.5, *g*_2_ = 0.8, and *R*_0_ = 2.5. Simulations begin with *I*_0_ = 100 initial infected seeds.

### 8 Hungarian contact data

For completeness, also in this case we explore the number of deaths estimated by our model under the two different scenarios when the population distribution and the mixing patterns are taken from real data. Panels *a* and *c* of Fig.S15 show the results for Scenario 1 while panels *b* and *d* of Fig.S15 refers to Scenario 2.

**Figure S15:**
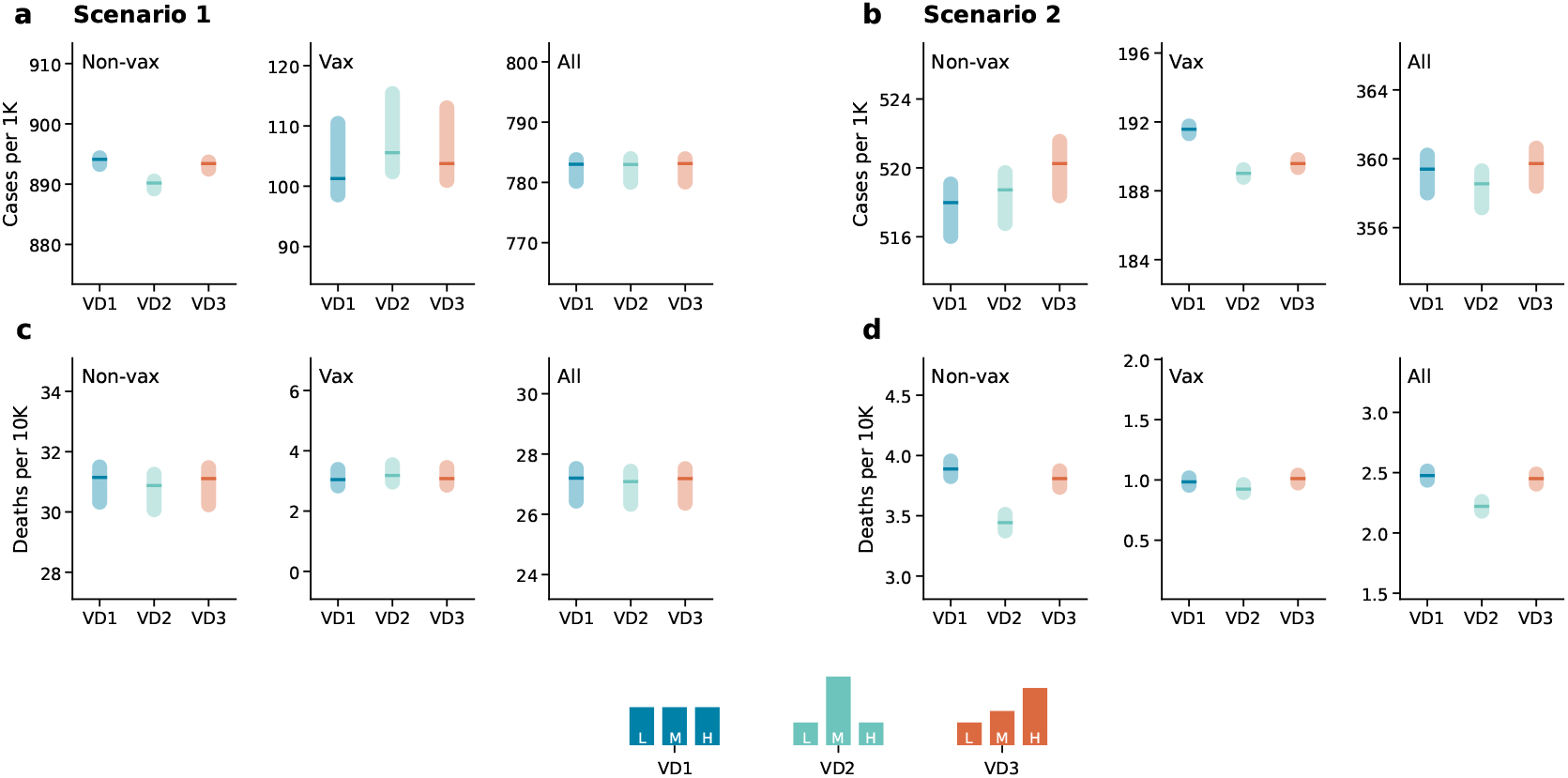
Epidemic outcomes under different vaccination distributions and real contact data.. Panels in *a* and *c* refer to Scenario 1 and display the number of cases per 1000 (panel *a*) and the number of deaths per 10000 (panel *c*), stratified by vaccination status (non-vaccinated, vaccinated, and overall) and under different vaccination distribution scenarios (*V D*0, *V D*2, *V D*3). Panels in *b* and *d* show the corresponding number for Scenario 2. Results refer to the median of 500 runs with confidence intervals. Epidemiological parameters: Γ = 0.25, Ψ = 0.4, *g*_1_ = 0.6 and *g*_2_ = 0.8. *R*_0_ = 2.7 for the third wave and *R*_0_ = 3 for the forth. Simulations start with *I*_0_ = 100 initial

Additionally, here we present the same results stratified by age and vaccination status. These figures explore the dynamics of infection and mortality under different vaccination distribution scenarios (*V D*1, *V D*2, and *V D*3).

**Figure S16:**
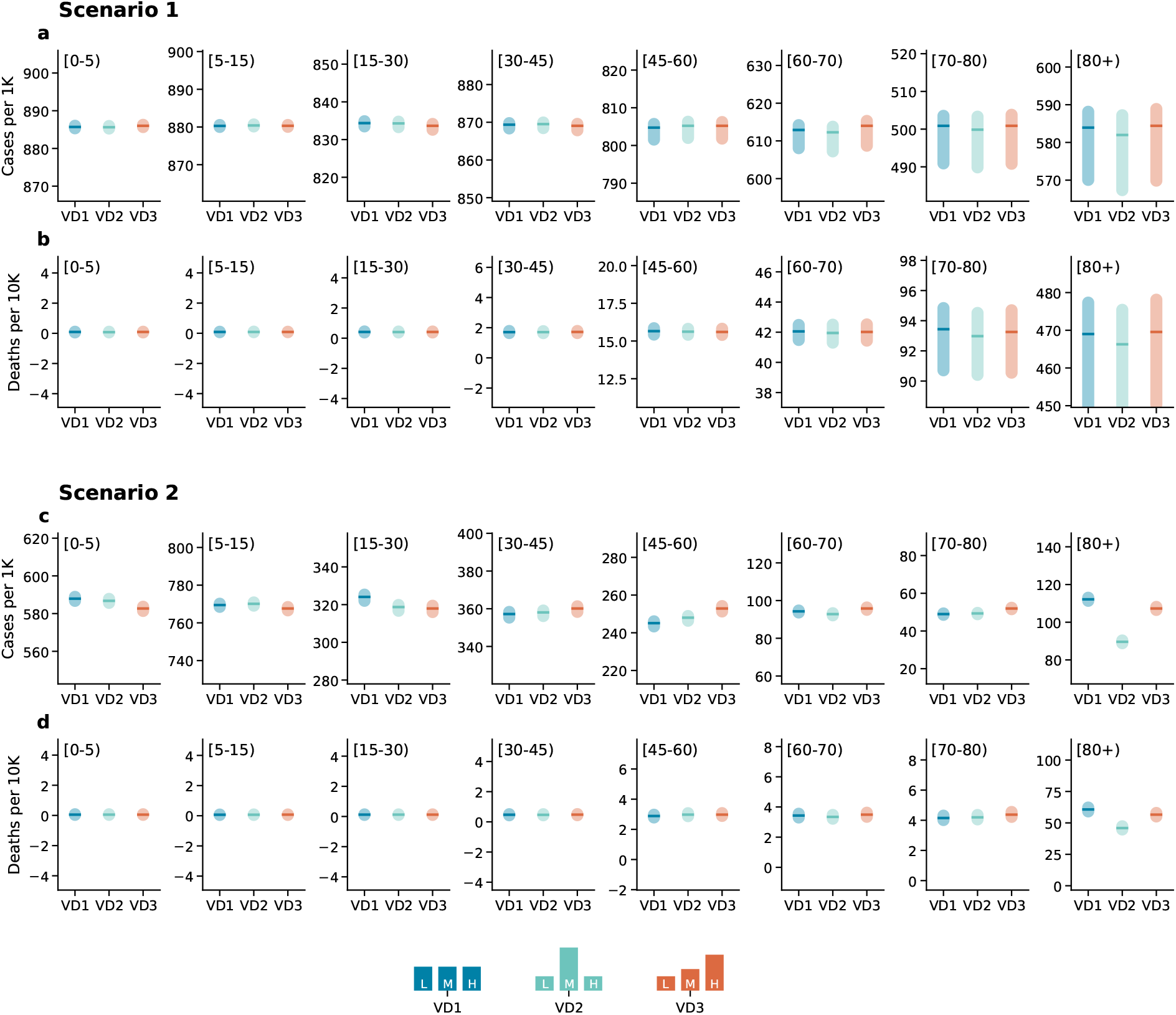
Epidemic outcomes under different vaccination distributions and real contact data, stratified by age for the entire population. Panels (*a*) and (*b*) refer to Scenario 1. Panel (*a*) displays the number of cases per 1000 individuals under different vaccination distributions (*V D*0, *V D*2, *V D*3), while panel (*b*) shows the corresponding number of deaths per 10, 000 individuals. Panels (*c*) and (*d*) present the same results for Scenario 2. Results represent the median of 500 simulations with confidence intervals. Epidemiological parameters: Γ = 0.25, Ψ = 0.4, *g*_1_ = 0.6, and *g*_2_ = 0.8. *R*_0_ = 2.7 for the third wave and *R*_0_ = 3 for the fourth. Simulations start with *I*_0_ = 100 initial infectious cases.

**Figure S17:**
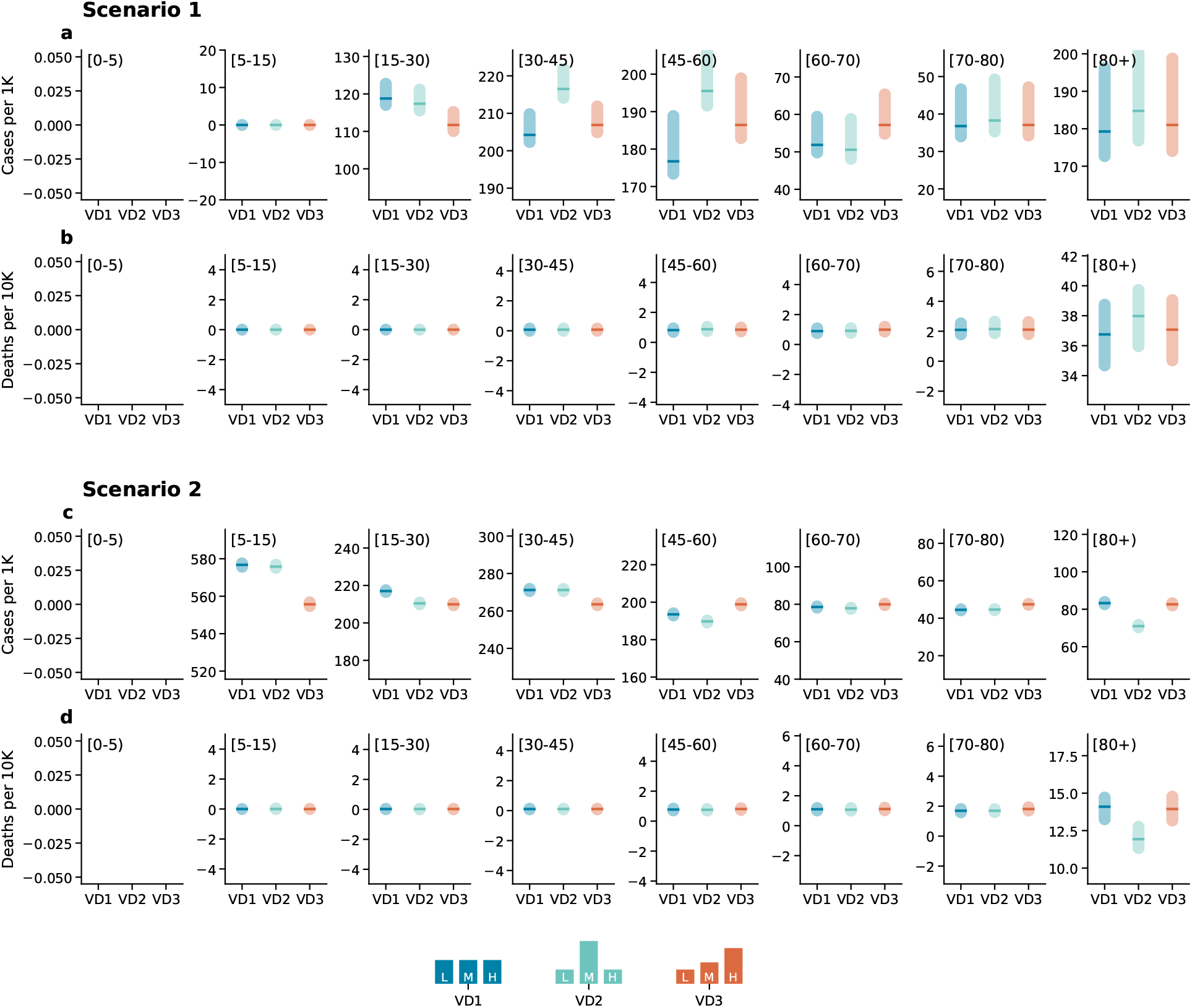
Epidemic outcomes under different vaccination distributions and real contact data, stratified by age for the vaccinated population. Panels (*a*) and (*b*) refer to Scenario 1. Panel (*a*) displays the number of cases per 1000 individuals under different vaccination distributions (*V D*0, *V D*2, *V D*3), while panel (*b*) shows the corresponding number of deaths per 10, 000 individuals. Panels (*c*) and (*d*) present the same results forScenario 2. Results represent the median of 500 simulations with confidence intervals. Epidemiological parameters: Γ = 0.25, Ψ = 0.4, *g*_1_ = 0.6, and *g*_2_ = 0.8. *R*_0_ = 2.7 for the third wave and *R*_0_ = 3 for the fourth. Simulations start with *I*_0_ = 100 initial infectious cases.

**Figure S18:**
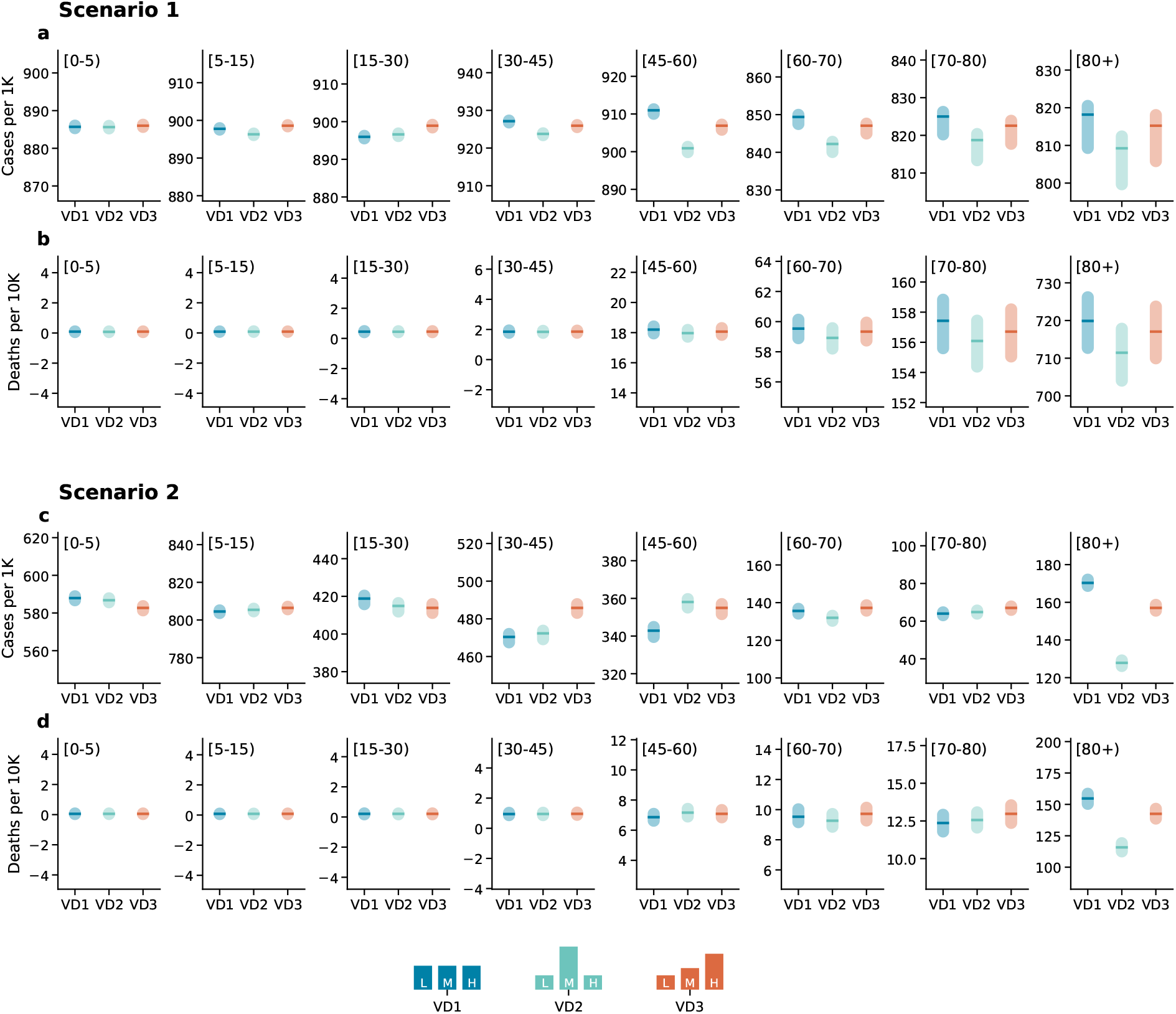
Epidemic outcomes under different vaccination distributions and real contact data, stratified by age for the non-vaccinated population. Panels (*a*) and (*b*) refer to Scenario Panel (*a*) displays the number of cases per 1000 individuals under different vaccination distributions (*V D*0, *V D*2, *V D*3), while panel (*b*) shows the corresponding number of deaths per 10, 000 individuals. Panels (*c*) and (*d*) present the same results Scenario 2. Results represent the median of 500 simulations with confidence intervals. Epidemiological parameters: Γ = 0.25, Ψ = 0.4, *g*_1_ = 0.6, and *g*_2_ = 0.8. *R*_0_ = 2.7 for the third wave and *R*_0_ = 3 for the fourth. Simulations start with *I*_0_ = 100 initial infectious cases.

## Footnotes

1 The *f* describes the a posteriori estimation of disease burden from traditional models featuring only age-structured contact matrices *C*_*ij*_

## Notes

### Competing Interest Statement

The authors have declared no competing interest.

### Author Declarations

The data collection adhered to European and Hungarian privacy regulations, approved by the Hungarian National Authority for Data Protection and Freedom of Information, as well as the Health Science Council Scientific and Research Ethics Committee (resolution number IV/3073-1/2021/EKU)

